# Advancing long-read nanopore genome assembly and accurate variant calling for rare disease detection

**DOI:** 10.1101/2024.08.22.24312327

**Authors:** Shloka Negi, Sarah L. Stenton, Seth I. Berger, Brandy McNulty, Ivo Violich, Joshua Gardner, Todd Hillaker, Sara M O’Rourke, Melanie C. O’Leary, Elizabeth Carbonell, Christina Austin-Tse, Gabrielle Lemire, Jillian Serrano, Brian Mangilog, Grace VanNoy, Mikhail Kolmogorov, Eric Vilain, Anne O’Donnell-Luria, Emmanuèle Délot, Karen H. Miga, Jean Monlong, Benedict Paten

## Abstract

More than 50% of families with suspected rare monogenic diseases remain unsolved after whole genome analysis by short read sequencing (SRS). Long-read sequencing (LRS) could help bridge this diagnostic gap by capturing variants inaccessible to SRS, facilitating long-range mapping and phasing, and providing haplotype-resolved methylation profiling. To evaluate LRS’s additional diagnostic yield, we sequenced a rare disease cohort of 98 samples, including 41 probands and some family members, using nanopore sequencing, achieving per sample ∼36x average coverage and 32 kilobase (kb) read N50 from a single flow cell. Our Napu pipeline generated assemblies, phased variants, and methylation calls. LRS covered, on average, coding exons in ∼280 genes and ∼5 known Mendelian disease genes that were not covered by SRS. In comparison to SRS, LRS detected additional rare, functionally annotated variants, including SVs and tandem repeats, and completely phased 87% of protein-coding genes. LRS detected additional *de novo* variants, and could be used to distinguish postzygotic mosaic variants from prezygotic *de novos*. Eleven probands were solved, with diverse underlying genetic causes including *de novo* and compound heterozygous variants, large-scale SVs, and epigenetic modifications. Our study demonstrates LRS’s potential to enhance diagnostic yield for rare monogenic diseases, implying utility in future clinical genomics workflows.

## Introduction

Despite extensive efforts to identify pathogenic variants using whole-genome protocols, approximately 50% of individuals with suspected rare genetic diseases remain undiagnosed (Graessner et al. 2021; Kingsmore et al. 2019; Costain et al. 2020). This diagnostic gap is not solely due to interpretative and clinical challenges— such as incomplete understanding of gene functions, genomic regions, and patient phenotypes that hinder accurate genotype-phenotype inference—but also partly due to technical limitations in the short-read sequencing (SRS) that is widely used for genome sequencing (Wojcik et al. 2023). For example, pathogenic variants may reside in parts of the genome that are "inaccessible" to SRS techniques. These regions consist of combinations of highly similar or highly repetitive sequences to which short reads (150-250 bps of typically paired-end sequences) are frequently unable to unambiguously map, thereby hindering variant detection. There are more than a thousand genes associated with these regions, some of which are known to be clinically relevant (Nurk et al. 2022; Wagner et al. 2022).

Similarly, structural variants (SVs) have been shown to contribute to many rare diseases (Groza et al. 2024; Merker et al. 2018), but the complexity of assembling and detecting SVs with conventional SRS approaches is well-documented (Treangen and Salzberg 2012; Olson et al. 2023). Numerous studies highlight high incidence of false positives and failure to detect SVs in low-complexity regions, such as those consisting of GC- or AT-rich repeats, tandem repeats, and in genomic loci characterized by segmental duplications (frequently long, and sometimes highly similar sequence copies), which are hotspots for SVs and experience some of the highest mutation rates, in both germline and soma (Alkan, Sajjadian, and Eichler 2011; Mills et al. 2011; Zook et al. 2020; Sudmant et al. 2015; Hodgkinson, Chen, and Eyre-Walker 2012).

In contrast, long-read sequencing (LRS) produces reads that are frequently two to three orders of magnitude longer than SRS. Such reads can accurately map across many large repetitive regions, thereby improving the detection of single nucleotide variants (SNVs) in homologous gene copies (Vollger et al. 2022) and facilitating *de novo* assemblies for more effective detection and evaluation of SVs (Cheng et al. 2021). Beyond improving variant resolution and detection, LRS offers long-range read-based haplotype phasing of variants, which can be critical for detecting compound heterozygous diagnoses, particularly when parental samples are unavailable to determine variant phase (Olivucci et al. 2024). Furthermore, most LRS uses single-molecule sequencing, facilitating the simultaneous detection of base modifications natively from the sequencing data and allowing for the study of genomic variation in conjunction with the methylation status of CpG sites (Wang et al. 2021).

Despite the reported advantages of LRS, short-read protocols are the standard-of-care diagnostic tests for genome sequencing due to well-established analysis protocols and pipelines, and the availability of large reference population databases for the prioritization of rare variants (Conlin et al. 2022). Though not yet widely clinically available, recent studies have demonstrated that LRS can identify causal variants in both known and novel disease-causing genes that had eluded detection by SRS, such as complex SVs and low complexity repeat expansions, in addition to facilitating phasing of variants also detected by SRS (Hiatt et al. 2024; Miller et al. 2021; Cohen et al. 2022; Lecoquierre et al. 2023; Ohori et al. 2023; Tayoun et al. 2024; Steyaert et al. 2024; X. Chen et al. 2023). However, many of these studies applied targeted LRS (adaptive sequencing) to loci of high phenotypic interest or included only a small number of samples. As a result, the additional diagnostic yield of genome-wide LRS over SRS is not clearly quantified at this time.

The two widely used, contemporary LRS technologies, Pacific Bioscience’s (PacBio) Hifi sequencing and Oxford Nanopore Technology’s (ONT) nanopore sequencing, differ fundamentally in their data generation methods, leading to differences in read lengths and error rates (Logsdon, Vollger, and Eichler 2020). PacBio HiFi reads range from 15-20 kilobases (kb) (Hon et al. 2020), while ONT long reads range from 10-100 kb, with ultra-long reads reaching 100-300 kb (Logsdon, Vollger, and Eichler 2020). ONT sequencing currently has lower read accuracy compared to PacBio HiFi and is subject to more frequent base-calling errors in homopolymers and short tandem repeats that lead to larger numbers of short insertion or deletion (indel) errors involving these sequences.

With continuous updates to pore chemistries and basecalling algorithms, ONT is improving in accuracy, making LRS on the ONT platform increasingly viable for clinical research (Mastrorosa, Miller, and Eichler 2023). ONT sequencing relative to other sequencing technologies can also be integrated into very rapid sequencing protocols to enable variant detection and characterization in time critical applications, such as acute care (Goenka et al. 2022; Gorzynski et al. 2022). In a recent study, Kolmogorov et al. 2023 demonstrated that state-of-the-art small and structural variant calling performance is achievable using ONT reads from a single flow cell at high throughput, including achieving a higher overall F1 score relative to Illumina for detecting SNVs according to Genome in a Bottle (GIAB) benchmarks (Olson et al. 2022). This protocol is potentially useful in diagnostic testing as it enables accurate *de novo* assembly and the combined evaluation of different alteration types (i.e., methylation, SNVs, small indels, and SVs) in a single analysis, avoiding the sequential application of multiple tests and offering a potentially significant advantage in terms of cost and simplicity relative to less comprehensive approaches.

Here we demonstrate the capabilities of this protocol to sequence and analyze rare disease samples, generating *de novo* assembly, haplotype-resolved small and SV calls, and haplotype-specific CpG methylation calls in a single workflow run. We applied the accompanying computational protocol, Napu (Nanopore Analysis Pipeline for U) (Kolmogorov et al. 2023) to a cohort that included trios, comprising healthy parents of children with undiagnosed rare diseases and several singletons for which parental data were unavailable. To test the ability of the method to detect known SVs of clinical significance, some samples with translocations or variants in segmental duplications were included, however most affected individuals had remained undiagnosed using Illumina SRS. We evaluated the clinical utility of our one-flow cell LRS-derived measurements by conducting a thorough comparison with those derived from SRS to identify the additional information yield LRS can provide. Out of 42 affected individuals in the cohort, LRS identified all the known diagnoses, demonstrated that a variant of unknown significance resulted in the known episignature of a rare syndrome and established or confirmed a diagnosis for 11 samples.

## Results

### Scalable ONT sequencing of blood samples yields high throughput and long read lengths

Several recent studies utilized ultra-long (≥100 kb) ONT sequencing to produce high-quality *de novo* assemblies of human genomes. However, multiple flow cells were used to achieve sufficient genomic coverage, as ultra-long DNA preparation protocols typically see lower sequencing yields. In our work, we further optimized the scalable Napu DNA processing and library preparation protocol for the ONT R10 chemistry (Kolmogorov et al. 2023) (methods), aiming to achieve sufficient coverage and read length to be useful for clinical sequencing. For 21 samples that were sequenced while we were still optimizing the protocol, we generated data from two R10.4.1 PromethION flow cells to reach the target throughput of >=30x coverage and >=30 kb read N50. For the remainder, each sample was sequenced with a single flow cell (average ∼113 gigabases (Gb) output per flow cell, corresponding to ∼36x coverage; average read length N50 of ∼32 kb; **Figure 1; Supplementary Table 1, Supplementary Figure 1)**. In total, we successfully sequenced 98 blood samples, comprising 26 trios (affected child plus unaffected parents), one quad (two affected siblings plus unaffected parents), one dyad (affected child plus single unaffected parent), a single unaffected mother of a proband, and 13 affected singletons. The majority of these samples were obtained from whole blood, while some were sourced from white blood cells (WBCs) and high molecular weight DNA. The median read identity aligned to the GRCh38 reference genome was 99.22%.

**Figure 1:**
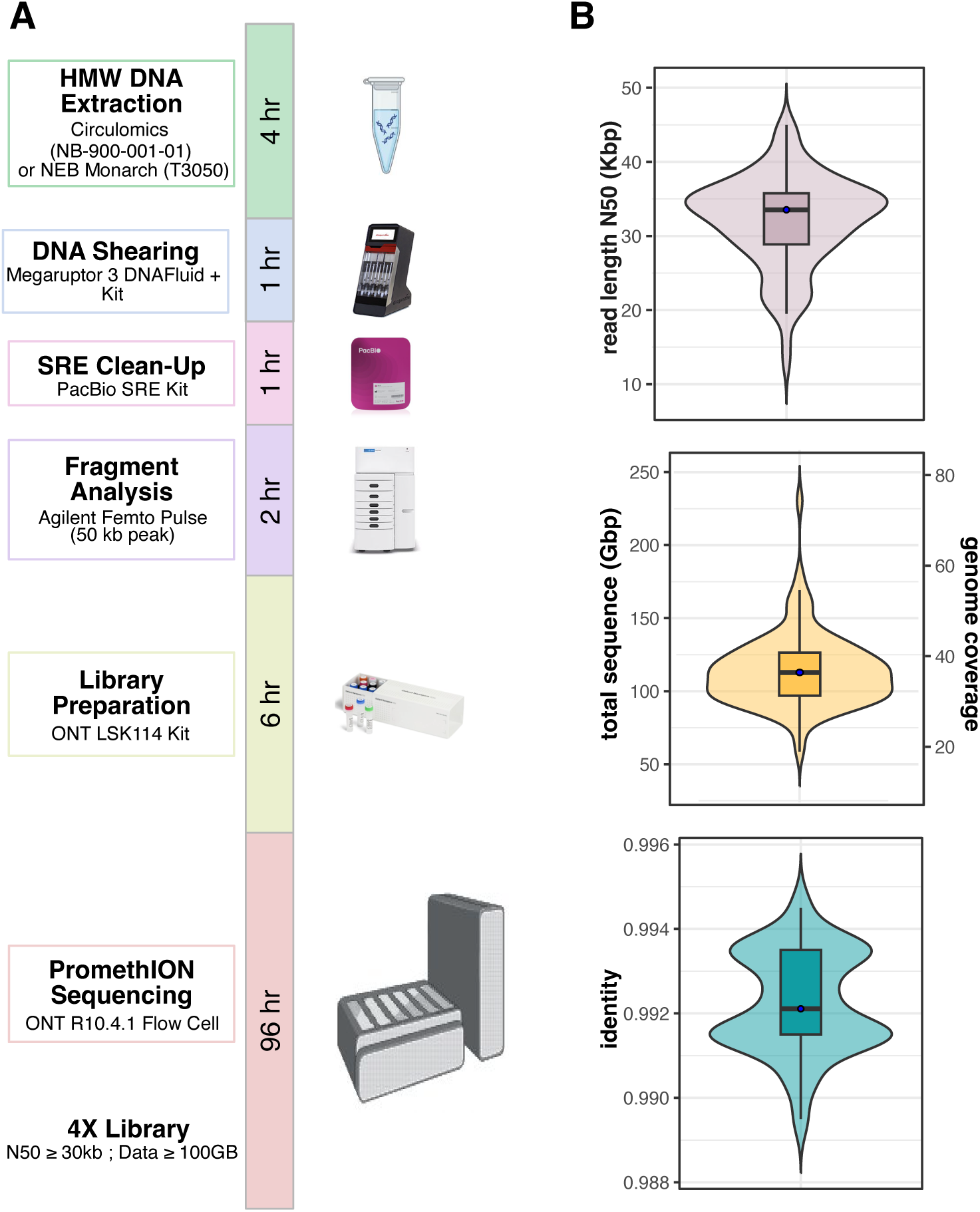
Single flow-cell scalable sequencing protocol. **A)** Cost-efficient, scalable, one-flow cell nanopore sequencing protocol. **B)** From top to bottom, read length N50, that is, the read length (y axis) such that reads of this length or longer represent 50% of the total sequence. Total sequenced bases/haploid human genome coverage (assuming a 3.1-Gbp genome) from total reads for each sample. Distribution of read identities (percentage of matching bases in reads when aligned to the reference genome) when aligned to T2T-CHM13 v.2.0. Supporting data are available in **Supplementary Table 1**.

### Genome completeness analysis reveals complex Mendelian disease genes well-covered by LRS only

Using GRCh38-aligned reads with a mapping quality of at least 10 (estimated 90% probability of correct mapping), the autosomal coverage across 21 affected individuals (20 probands, 1 affected sibling), for which both SRS and LRS data was available, was similar for LRS (median 35.21, minimum 24.26, maximum 46.91) and SRS (median 33.52, minimum 23.33, maximum 45.25) (**Supplementary Table 2)**. We quantified the percentage of callable bases (coverage between 10x to 80x) in the reference genome that we could call variants against (methods). LRS showed a higher proportion of callable bases (average 0.82% more) than SRS, covering 92.18% of the GRCh38 reference genome; LRS also had more high coverage and fewer low or no coverage bases than SRS **(Figure 2A, Supplementary Table 3).** Mapping to T2T-CHM13, the autosomal coverage increased for all of these samples (LRS median 37.17x, SRS median 36.8x). Reinforcing the study by Aganezov et al., (Aganezov et al. 2022) relative to GRCh38, a higher percentage (5.27%) of the T2T- CHM13 genome was callable by LRS (93.99%) vs. SRS (88.27%) **(Figure 2A, Supplementary Table 4-5)**, indicating the benefits of reference-based LRS analysis will grow with a transition to more complete telomere- to-telomere (T2T) and pangenome references (Liao et al. 2023). With GRCh38-aligned reads, we found many >1 kb regions exclusively callable by LRS consistently across all chromosomes (**Figure 2B, left** showing proband RGP_696_3 as a representative example). Interestingly, the relatively few SRS-only callable regions all went away on mapping to the T2T-CHM13 reference **(Figure 2B, right),** with manual analysis confirming they were the result of mapping artifacts caused by the incompleteness of GRCh38.

**Figure 2:**
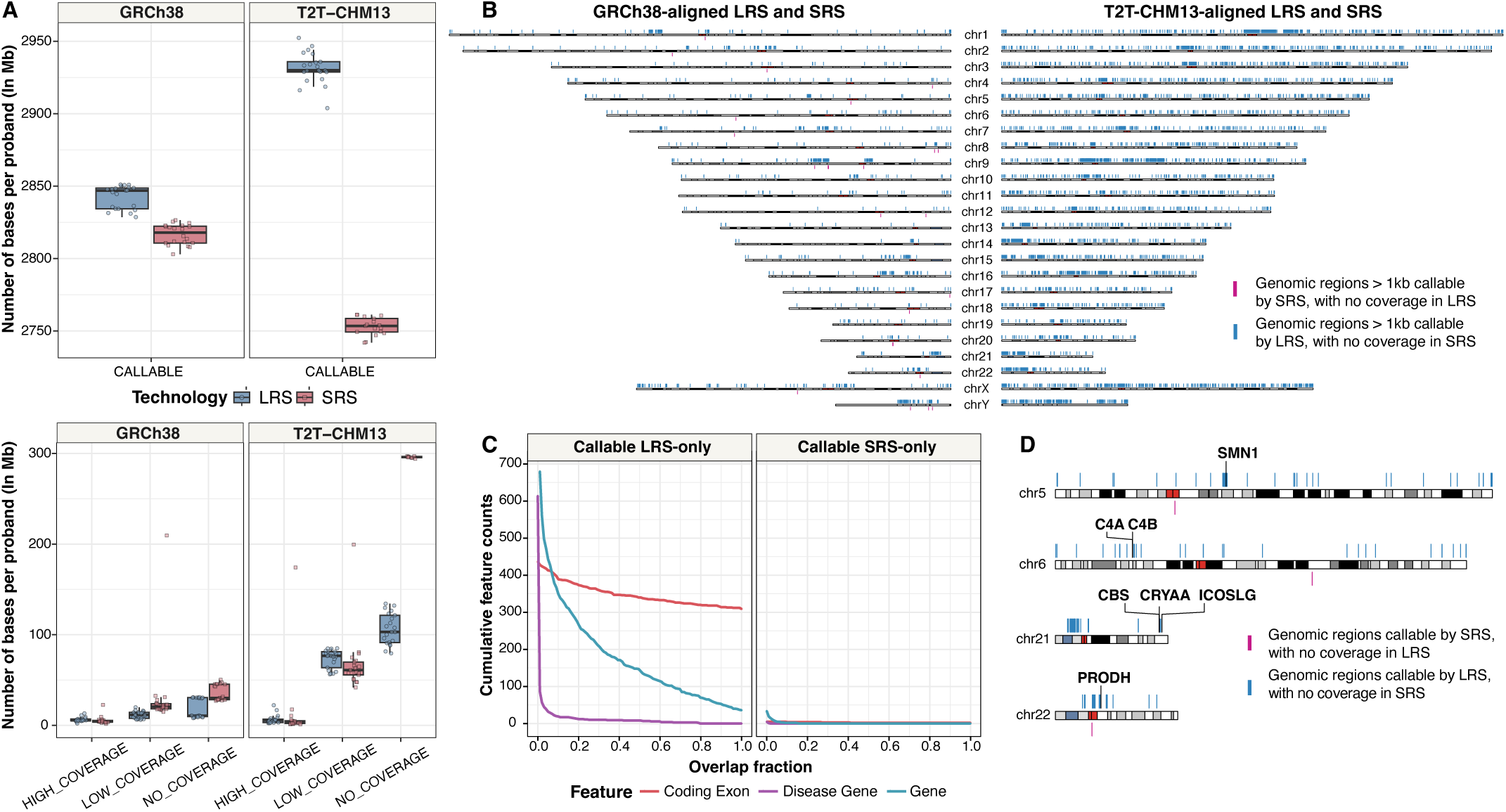
Genome completeness analysis reveals complex Mendelian disease genes callable by long-read sequencing only. **A)** Genome-wide coverage distribution across all probands for GRCh38 and T2T-CHM13, calculated using reads with a mapping quality (MAPQ) greater than 10. Assembly gaps and simulated centromeric regions were excluded for GRCh38-aligned reads. Bases are categorized into four coverage levels: CALLABLE (10-80x), HIGH_COVERAGE (>80x), LOW_COVERAGE (0-10x), and NO_COVERAGE (0x). For proband RGP_696_3, **B)** Ideogram for GRCh38 (left) and T2T-CHM13 (right), showing genomic regions > 1 kb callable by LRS with no coverage in SRS (blue) and regions > 1kb callable by SRS with no coverage in LRS (magenta). Red cytoband represents the centromere. **C)** Cumulative counts of genomic features (coding exons, Mendelian disease genes and all protein-coding genes) based on overlap fraction. The x-axis shows the fraction of each feature’s length. The y-axis shows the number of genomic features with LRS-only/SRS-only callable coverage over at least a fraction (x) of their length (y-axis limit is set to 700 for clarity). **D)** Ideogram highlighting seven Mendelian disease genes that have most of their length callable by LRS-only. Red cytoband represents the centromere. B,C,D is shown for one proband, and supporting data for other probands are available in **Supplementary Table 2**.

A median of 111 genes per affected individual had at least half their length covered by LRS and not by SRS, including a median of 5 Mendelian disease genes (**Supplementary Table 6**); 38 genes had their entire length callable by LRS alone, and 280 genes had at least one coding exon per gene that was entirely covered by LRS alone, including the first coding exon in ∼134 of these genes, where SRS is often known to exhibit reduced coverage due to high GC content (Porreca et al. 2007; Hoppman-Chaney et al. 2010; Hu et al. 2009; Valencia et al. 2012). Reversing the analysis, SRS exclusively covered a median of 2 coding exons, all of which appeared to result from mismapping in segmental duplications, and did not fully cover any genes that LRS did not. Examining one affected individual as an example, seven Mendelian disease genes (*SMN1, C4A, C4B, CBS, CRYAA, ICOSLG,* and *PRODH*) had at least half their length covered by LRS and not by SRS (**Figure 2C and 2D)**; *C4A* and *C4B*, associated with complement component 4a and 4b deficiency, are located within the highly polymorphic MHC locus and overlap regions of high-identity segmental duplications (Awdeh and Alper 1980) **(Supplementary Figure 2)**. Similarly, *SMN1,* with >99.9% sequence identity to its paralog *SMN2*, resides in a large and complex segmental duplication region (X. Chen et al. 2023). These results highlight the added value of LRS in capturing genetic information that may be missed by traditional short- read methods.

### LRS detects additional rare functionally annotated small variants

To carry out an unbiased comparison of the number of functionally annotated variants (FAVs), we applied a clinical annotation pipeline (methods) to LRS and SRS small variant call sets (variants < 30 bps) for 21 affected individuals with data from both sequencing technologies. FAVs were defined as non-homozygous reference exonic variants, with predicted HIGH/MODERATE impact or loss of function. With genotype quality (GQ) >= 20, we observed a high concordance between functional annotated single nucleotide variants (FA-SNVs) identified by SRS and LRS, with 91.6% of LRS FA-SNVs being called by SRS and 97.5% of SRS FA-SNVs being called by LRS **(Figure 3A)**. LRS detected a median of 1437 FAVs not detected by SRS per sample, whereas SRS called a median of 494 FAVs not detected by LRS **(Figure 3B).**

**Figure 3:**
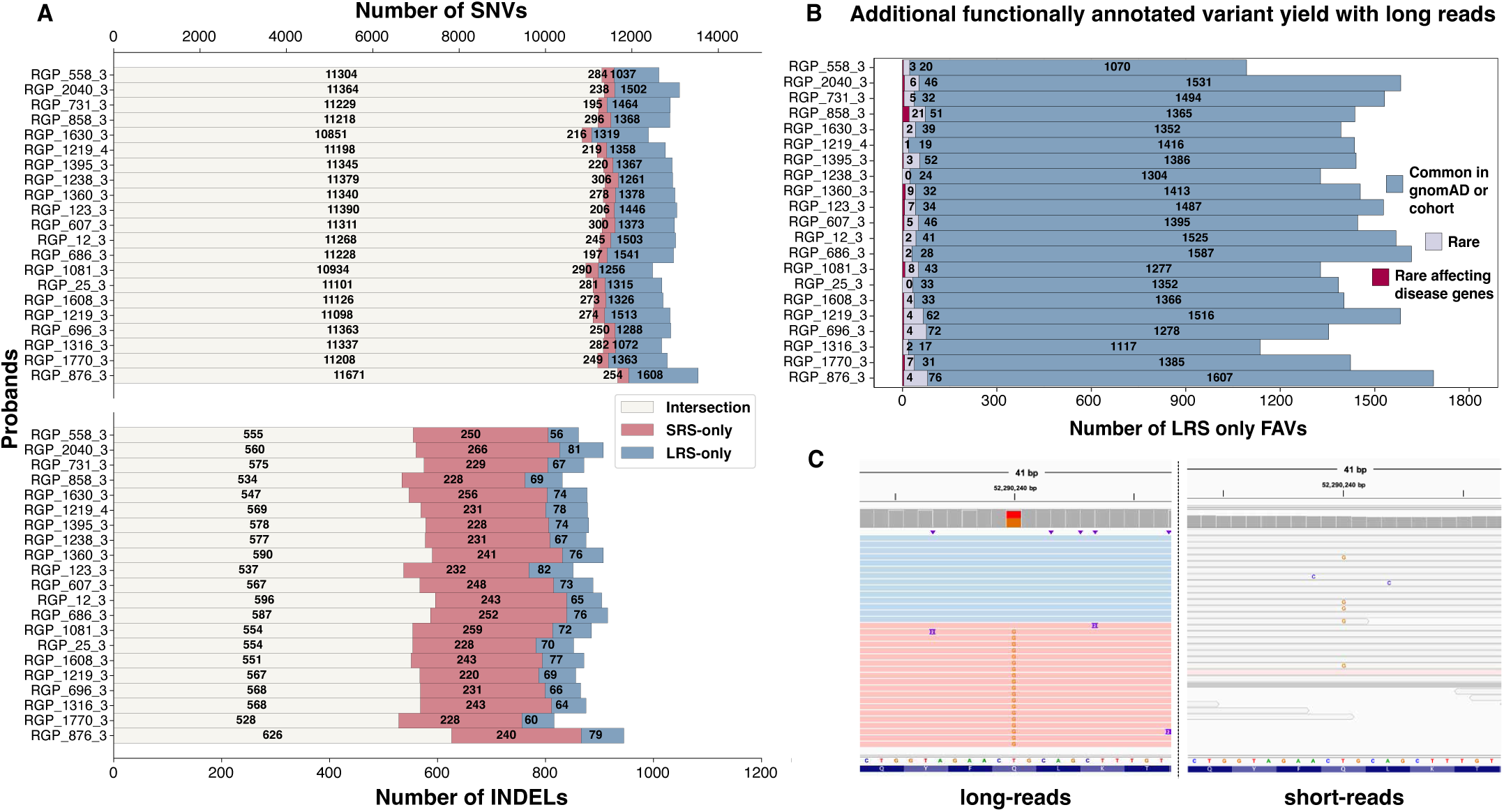
LRS detects additional rare functionally annotated small variants. **A)** Comparison of functionally annotated HIGH and MODERATE impact functionally annotated small variants (top - SNVs, bottom - indels) between SRS and LRS. **B)** Linear breakdown of LRS-only FAVs reveals additional rare variants in Mendelian disease genes. **C)** Example from sample RGP_1081_3 showing a rare, heterozygous, MODERATE impact missense variant in *KRT86*, a Mendelian autosomal dominant gene associated with monilethrix (OMIM#158000), located in a region unmappable with short-reads (note this was not found to be clinically relevant in the proband).

Stratification revealed that per sample, a median of 41 LRS-exclusive FAVs were rare (allele frequency <0.001 in gnomAD v3 and unique to a single family; 38 FA-SNVs, 4 FA-indels per sample), of which a median of 4 were located in Mendelian disease genes. Further, 80% of LRS-exclusive rare FAVs overlapped segmental duplications, and 66% were in low short-read mappable regions (sourced from GIAB GRCh38 stratifications). Manual investigation confirmed that SRS maps poorly to these sites (**Figure 3C** provides an example), highlighting the additional yield of LRS in SRS-inaccessible regions.

Conversely, of SRS-exclusive FAVs, a median per sample of 24 were rare (20 FA-SNVs, 4 FA-indels), of which 4 affected Mendelian disease genes **(Supplementary Figure 3A)**. More than 200 of the SRS exclusive FAVs per sample were indels; however, notably, LRS and SRS called similar median numbers (n=4) of rare exclusive FA-indels, despite indel calling being a challenge with ONT in homopolymers and tandem repeats. A median of 46% of SRS-exclusive rare FAVs overlapped segmental duplications or were in low short-read mappable regions **(Supplementary Figure 3B)**. A manual investigation of 21 variants in these regions (one randomly picked from each sample) revealed 16 likely false positives (multi-allelic or allele balance<0.2). However, most SRS-only FAVs were called by LRS but with slightly lower GQ. Since GQ calibration between SRS and LRS variant callers is different, we compared unfiltered call sets and found median per sample LRS-only FAVs doubled to 3046 (with 99 rare (90 FA-SNVs, 13 FA-indels) of which 15 were in Mendelian disease genes), while SRS-only FAVs dropped to 334 (with 22 rare (16 FA-SNVs, 2 FA-indels) of which 5 were in Mendelian disease genes), suggesting that future technology development leading to more confident, higher GQ LRS variant call sets could enhance the benefits of LRS over SRS for FAV detection **(Supplementary Figure 4)**.

### Comparison of *de novo* SNVs between SRS and LRS identifies postzygotic mosaicism

Across 21 affected individuals with both LRS and SRS data, after strict filtering (methods) and restricting to *de novo* SNVs (DN-SNVs), genome-wide LRS detected a median of 65 per sample, while SRS detected 62. **(Figure 4A; Supplementary Table 7**). Annotated DN-SNVs (methods) were also similar for LRS (median 47) and SRS (median 45). Aggregating across samples, 972 (74.6% of SRS DN-SNVs and 74.26% of LRS DN-SNVs) whole-genome DN-SNVs and 689 (74.25% of SRS DN-SNVs and 76.73% of LRS DN-SNVs) annotated DN-SNVs, respectively, were called by both technologies **(Figure 4B)**.

**Figure 4:**
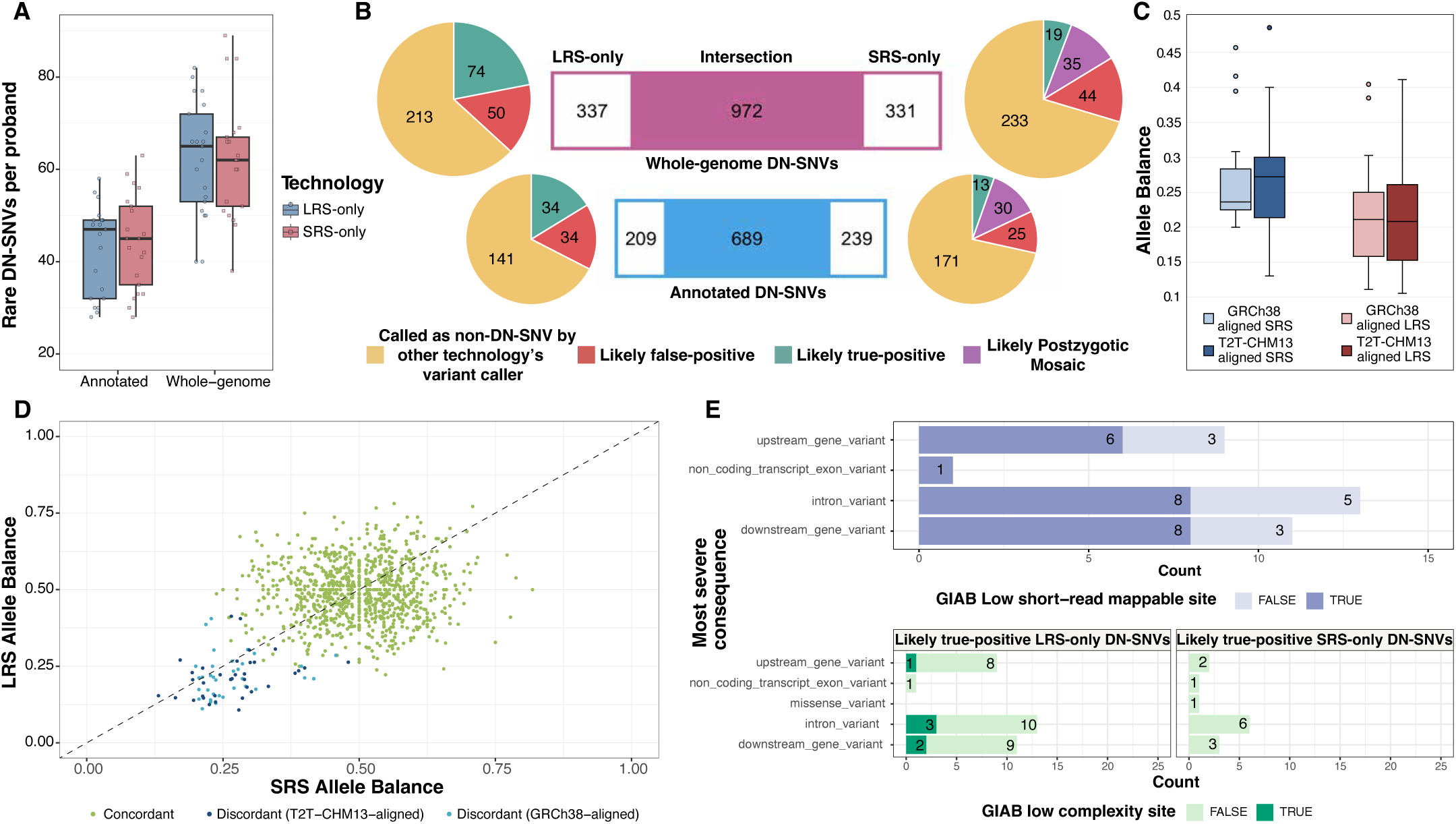
De novo SNV comparison between SRS and LRS. **A)** Counts of rare DN-SNVs (genome-wide and annotated) called exclusively by each technology (LRS or SRS). **B)** Comparison between LRS and SRS DN-SNV callsets. Bar charts in the center represent concordance. Pie charts on each side (left for LRS-only and right for SRS-only) show the proportion of exclusive DN-SNVs that upon IGV inspection are found to be likely false-positive, called by the other technology as non-DN-SNV, likely true-positive, or likely postzygotic mosaic. **C)** Allele-balance of likely postzygotic mosaic DN-SNVs in SRS (left) and LRS (right) reads, mapped to both GRCh38 and T2T-CHM13. **D)** Correlation of allele-balance between SRS and LRS for potential mosaic DN-SNVs compared to the concordant set (DN-SNVs called by both technologies). Allele-balance is consistent across GRCh38 and T2T-CHM13 mapped reads. **E)** (Top) Likely true-positive LRS-only DN-SNVs in GIAB low short-read mappable regions. (Bottom) Likely true-positive LRS-only and SRS-only DN-SNVs stratified by overlap with GIAB low-complexity regions. Supporting data is in **Supplementary Table 7-14**.

Sequencing technology-specific DN-SNVs were categorized into 4 validation categories - “A - Called as non- DNV by other technology’s variant caller”, “B - Likely false-positive”, “C - Likely postzygotic mosaic” and “D - Likely true-positive” (**Figure 4B, Supplementary Table 8-9;** methods). Most sequencing technology- specific DN-SNVs fell into category A, as they were called by the other technology’s variant caller, but were subsequently filtered out as a potential *de novo* variant due to being either present in one or both parents or being common within the cohort **(Supplementary Table 10-13)**.

Excluding the variants in categories A and B, we examined the remaining 54 SRS-only DN-SNVs and found LRS read evidence for 50, though they were not genotyped by DeepVariant. Upon investigation, we consistently found the alternate allele phased to one haplotype in LRS, but many reads on that haplotype showed the reference allele **(Supplementary Figure 5)**. Possible reasons included incorrect phasing, copy number variation, and mosaicism. Normal coverage and consistent phasing with neighboring SNVs ruled out the first two. The allele balance (AB) was consistent when mapping LRS and SRS to either GRCh38 or T2T- CHM13, suggesting it was not caused by an obvious read-reference mapping artifact **(Figure 4C, Supplementary Figure 6)**. LRS AB showed overall better consistency than SRS, potentially due to better mapping to both GRCh38 and T2T-CHM13. Using stringent filters (methods), we classified 35/54 of these discordant DN-SNVs (and 25/38 annotated DN-SNVs) as likely postzygotic mosaics, confirmed by SRS and LRS **(Supplementary Table 14)**. Convincingly, these likely postzygotic mosaics had lower AB compared to the concordant DN-SNVs (i.e. DN-SNVs called by both SRS and LRS) **(Figure 4D)**. As future work, integration of a somatic long-read variant caller into our pipeline should make it possible to confidently call these variants with LRS and identify them as postzygotic mosaic, which is often difficult with SRS due to the absence of read-based phasing information for many variants.

After this analysis, there were only 19 whole-genome and 13 annotated likely-true positive, non-postzygotic mosaic SRS-exclusive DN-SNVs (5.7% and 5.4% respectively, of the total SRS-exclusive DN-SNV callset). Several of these SRS-only true-positive DN-SNVs were observed on LRS but were either multi-allelic or had very low base quality, in part due to underlying homopolymers and di-nucleotide repeats. However, some missed variants on LRS lack a clear explanation **(Supplementary Figure 7)**.

Conversely, all remaining LRS-only DN-SNVs were categorized as likely true-positives (74 genome-wide and 34 annotated), since none showed evidence of potential postzygotic mosaicism (AB evenly distributed around 0.45), as DeepVariant uses phasing information and is not trained to detect somatic variants **(Supplementary Figure 8)**; very few LRS-only DN-SNVs showed any SRS read evidence. LRS-only likely true-positive DN-SNVs were stratified by mappability and complexity using GIAB GRCh38 stratifications. 23/34 (68%) annotated LRS-only likely true-positive DN-SNVs (and 51/74 (69%) genome-wide) were in low short-read mappable regions **(Figure 4E (top), Supplementary Figure 9)**. Additionally, 6/34 LRS-only true- positive DN-SNVs were in low-complexity regions, compared to none of the 19 SRS-only true-positive DN- SNVs **(Figure 4E (bottom))**. Notably, we observed the presence of LRS-only DN-SNV clusters on specific chromosomes in a few samples **(Supplementary Figure 10)**. Some of these clusters overlapped segmental duplications but were retained in the callset as they were individually high-quality DN-SNVs with a lack of evidence on well-covered parental reads and unique within cohort. These could be representative of gene- conversion events or the mapping to copy-balanced alternative duplicon copies, but further analysis of surrounding structural variation is needed to determine the likelihood of these scenarios.

Reviewing the *de novo* variants in the 20 families analysed above, we were able to make new diagnoses for three affected individuals. In each case the variants were discovered by both SRS and LRS and prior interpretation of SRS had overlooked them. The diagnoses could now be made based upon recent findings reported in the literature, and evidence the benefits of reanalysis. In two families (RGP_607 and RGP_696) with a neurodevelopmental phenotype, a highly recurrent *de novo* single base insertion (chr12:120291839T>TA) in the non-coding gene *RNU4-2* was detected in both SRS and LRS. These cases contributed to the recent report of *RNU4-2* as a novel disease gene and frequent cause of syndromic neurodevelopmental delay (Y. Chen et al. 2024). In family RGP_123 with a neurodevelopmental phenotype, a *de novo* 5’UTR splice variant in the gene *GLUL* was prioritized in the LRS analysis. The variant was noted but not considered high priority at the time of SRS analysis due to lack of a compound heterozygous variant and limited phenotype overlap given the only Mendelian condition reported at the time was recessive. However, a novel *de novo* mechanism of disease and new phenotype-association of developmental and epileptic encephalopathy, in keeping with our proband, has recently been reported for *GLUL* (Jones et al. 2024). As the report included functional validation of this proband’s *GLUL* variant (chr1:182388752T>C) in a second unrelated proband, we now consider the variant causal.

### Accurate characterization of structural variants and tandem-repeat expansions with LRS

Comparing LRS SV calls (called by Hapdiff, similar results were found using Sniffles **(Supplementary Table 15-16)**) to SRS SV calls (called by GATK-SV, which includes, Wham, and depth-based algorithms (Collins et al. 2020)) revealed that LRS detected a median of 22,561 SVs (>= 50bp) per individual, compared to 16,496 SVs by SRS **(Supplementary Table 17)**. Since LRS variant caller reports duplications as long insertions, we combined LRS and SRS duplications as insertions for comparison. Per proband, LRS detected a median of 13,738 insertions and 41 inversions, approximately 2- and 4-times the medians identified by SRS (Insertions - 7131, Inversions - 10) **(Figure 5A)**. Deletion numbers were comparable; LRS detected a median of 8944 deletions, while SRS detected 9337.

**Figure 5:**
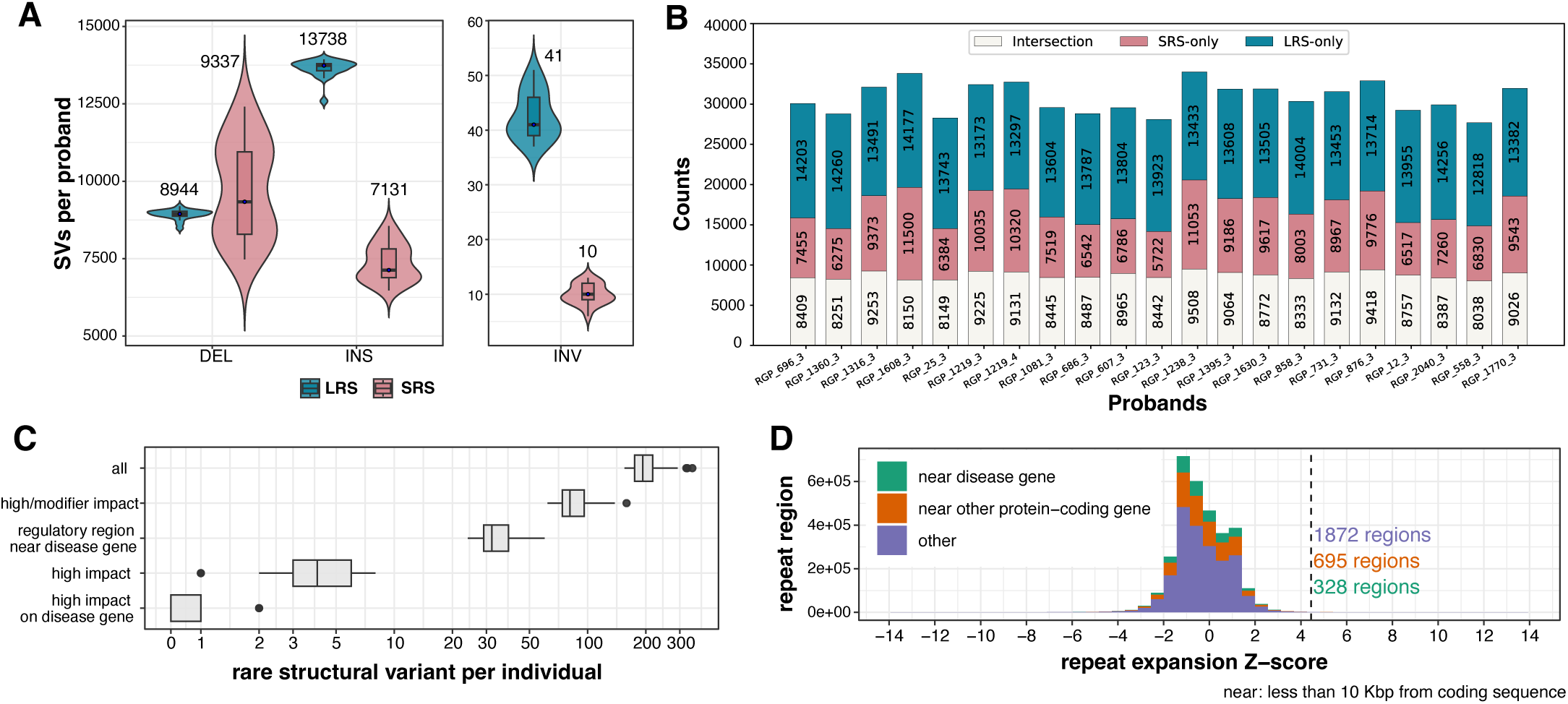
Characterization of structural variants and tandem-repeat expansions with LRS. **A)** Counts of LRS and SRS SVs (deletions, insertions and inversions) per proband. **B)** Comparison of LRS and SRS SVs using a fuzzy-matching approach implemented by *sveval* (Supporting data in Supplementary Table 18). **C)** Number of rare structural variants (allele frequency of 0.01 or less) in each individual with different profile. The violin plots represent the distribution across probands, the dots highlight the median values. The *high* and *modifier* impact prediction came from SnpEff (HIGH/MODIFIER impact classes). Regulatory regions are candidate cis-Regulatory Elements from ENCODE with enhancer-like signature. **D)** Expansion scores at annotated simple repeat sites across all probands. The vertical dotted line highlights repeats that are significantly expanded (adjusted P-value < 0.01 and fold-change > 2) compared to the controls. Regions at less than 10 Kbp of coding exons are highlighted in green (for known disease genes) and orange (for other protein-coding genes).

Stratifying by length, an assembly-based and a reference-based LRS variant caller (Hapdiff and Sniffles, respectively) showed consistent insertion counts across the length spectrum, detecting more insertions than SRS throughout **(Supplementary Figure 11)**. For deletions, LRS and SRS counts were comparable up to 100kb. Beyond this, more deletions were observed in SRS data, possibly influenced by false-positives or alignment-based LRS methods not being well calibrated for detecting split read mappings.

Intersecting the SV callsets, LRS called a median of 13,714 SVs per affected individual not called by SRS **(Figure 5B, Supplementary Table 18-19)**. Inversely, SRS called a median 8003 SVs not called by LRS. Manual investigation of a set of 10 randomly chosen SRS-only SVs confirmed at least 9 to be false positives, reinforcing earlier findings of high error rates **(Supplementary Table 20;** (Alkan, Sajjadian, and Eichler 2011; Mills et al. 2011; Zook et al. 2020; Sudmant et al. 2015; Hodgkinson, Chen, and Eyre-Walker 2012)). For example, for SRS-only deletions, we observed many cases without paired-end reads spanning the deletion, and showing uniform coverage throughout with LRS **(Supplementary Figure 12)**. We also found cases involving multiple overlapping fragmentary SRS SVs in regions where LRS had called a single sequence-resolved SV (**Supplementary Figure 13**). The high false-positive rate of the SRS SVs was also evident from a very high heterozygous to homozygous ratio (median 4.84, maximum 6.88), compared to median 1.67 for LRS SVs (for comparison, median het/hom for small variants; LRS - 1.65 and SRS - 1.63; **Supplementary Table 21-22)**. Conversely, examining a random set of 10 LRS-only SVs, none were false calls **(Supplementary Table 23, Supplementary Figure 14)**.

After annotating the LRS SVs with frequencies and functional impact predictions (methods), we found, on average per affected individual, 208 rare SVs (allele frequency below 0.01), 4 rare SVs with a high predicted impact (typically disrupting exonic sequences), and a rare SVs with a high predicted impact on a known disease gene in 29% of cases (**Figure 5C**), with one of these contributing to the patient’s phenotype and now considered diagnostic (case DSDTRN17 described below). We also identified, on average, 35 rare SVs overlapping a regulatory element of a known disease gene. SVs were also annotated with AnnotSV (Geoffroy et al. 2018), and we found at least one rare deletion with a ranking score higher than 0.9 (score adapted from ACMG/ClinGen’s recommendations and equivalent to “likely pathogenic”) in 6 affected individuals (∼15%).

In trios, we also identified, on average, 3.2 candidate *de novo* SVs by comparing the probands with their parents (methods), three of which overlapped coding sequences although not from a known disease gene.

We used the phased LRS SV calls within annotated simple repeats to measure the size variation at these sites for each haplotype. We then selected repeat expansions for further investigation if they were outliers in this cohort and compared them to a set of control high-quality phased *de novo* assemblies from the Human Pangenome Reference Consortium (Liao et al. 2023) and the Human Genome Structural Variation Consortium (Ebert et al. 2021) **(Figure 5D)**. On average, per proband, 84 simple repeat sites were significantly expanded compared to controls (adjusted P-value < 0.01 and fold-change > 2). 29.5 sites were, on average, located at less than 10 Kbp from a protein-coding gene, 9.2 of which were from a known disease gene. Of note, 28 expanded repeat sites in 26 probands overlapped directly with coding sequences. A clinical review of these variants did not allow us to confidently implicate any with a participant’s phenotype.

### Base-level characterization of sex chromosomal translocations

Translocations between sex chromosomes are challenging to detect because of their representation in the GRCh38 reference genome and sequence similarity that may confuse read mapping and SV detection tools. Two patients in our cohort had XX male syndrome caused by a translocation between the chrX and chrY p- arms. Although some evidence could be found manually, these variants could not be detected by SV detection tools using SRS data. Both translocations had been detected with optical mapping from BioNano (Sahajpal et al. 2021). In our long-read dataset, we were able to discover the breakpoints of each translocation supported by multiple split-mapped reads when mapped to the T2T-CHM13 reference genome **(Supplementary Figure 15A-B)**. The breakpoints were consistent with the copy number changes inferred from read coverage. Of note, one translocation was detected by Sniffles2 from reads mapped with NGMLR, a more sensitive and SV- oriented long-read mapper. In comparison to other technologies, long-reads from ONT allowed us to detect the presence of the translocations and precisely predict their breakpoints’ locations.

### Detecting gene fusion and conversion of *CYP21A2*

The *CYP21A2* gene, responsible for 21-hydroxylase deficiency, is located in a tandemly-duplicated segmental duplication that is about 30 kbp long. The most common pathogenic variants arise from gene conversion event with the *CYP21A1P* pseudogene located in the upstream module, or deletions that create CYP21A1P-CYP21A2 fusions (Merke and Auchus 2020). Although the Napu pipeline detected some pathogenic variants, we developed a specialized tool to fully characterize this region at the haplotype level (see Methods, manuscript in preparation). Using this new approach, we identified compound heterozygous pathogenic variants for all four probands with 21-hydroxylase deficiency in our cohort **(Supplementary Figure 15C)**. Three cases involved a pathogenic SNV and a CYP21A1P-CYP21A2 fusion. Of note, two cases were particularly challenging to analyze because they carried a haplotype with three modules (two with the pseudogene and one with a gene-converted gene). Overall, our approach was able to identify and phase the pathogenic alleles in all probands and showed perfect Mendelian consistency with their parents.

### Phasing with long reads reveals compound heterozygous variants

Napu generates harmonized and phased structural and small variant calls, providing a comprehensive representation of sample variants. Across all samples, the median phase block NG50 was 2.16 Mb **(Supplementary Figure 16)**. This phased view helps characterize complex regions containing multiple variants on both haplotypes to identify compound heterozygous variants in genes associated with Mendelian recessive disorders. Among protein-coding genes, a median of 17,365 (87%) out of 20,048 per individual were entirely within a single phase block (**Figure 6A, Supplementary Figure 17)**.

**Figure 6:**
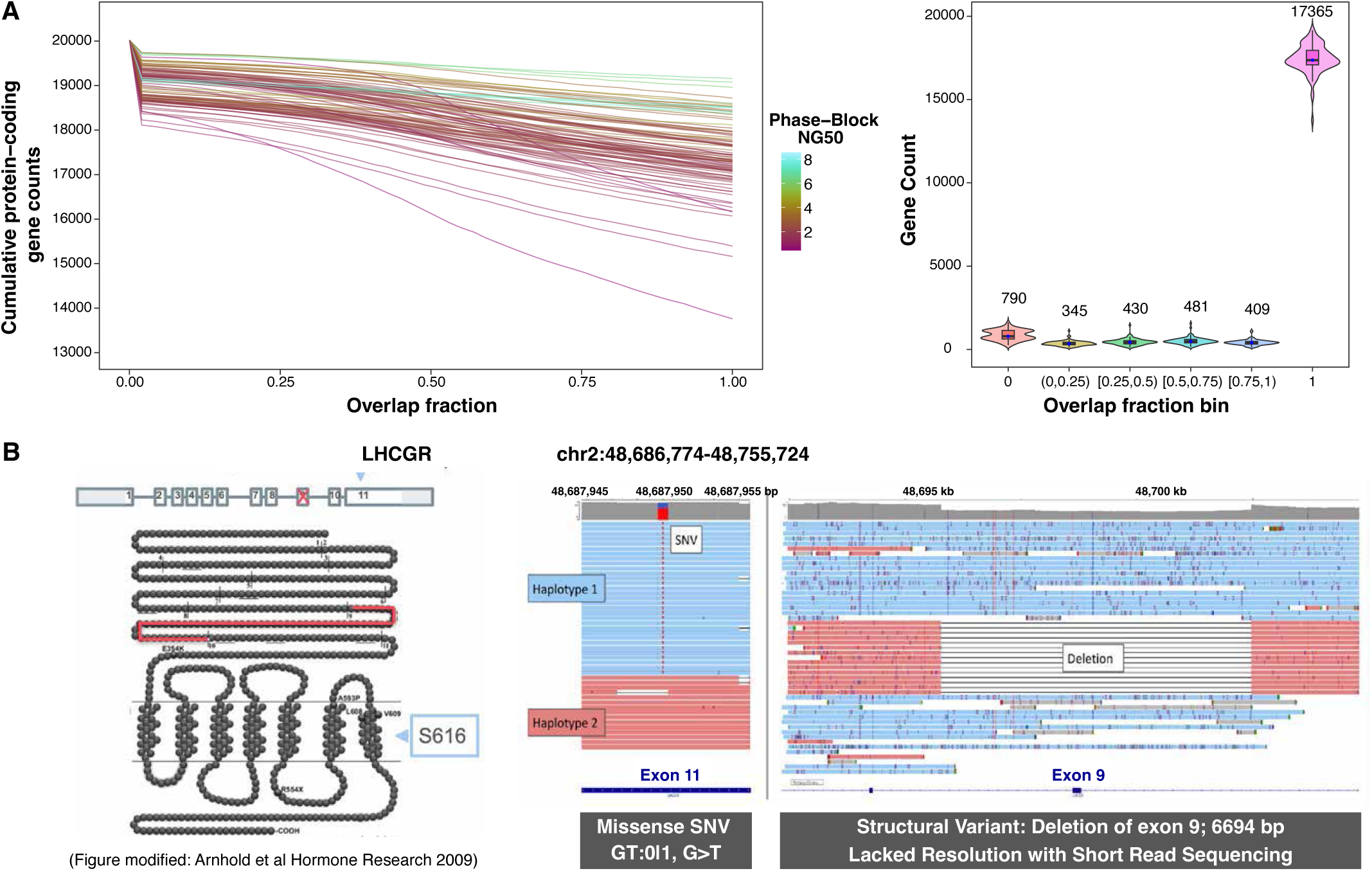
Phasing with long reads reveals compound heterozygous variants in protein-coding genes. **A)** First plot shows cumulative counts of protein-coding genes overlapping a single phase-block with varying overlap fractions. The x-axis shows the fraction of each gene’s length. The y-axis represents the number of genes that are at least x fraction phased by a single phase-block. Each line corresponds to a sample and is colored by its phase-block NG50. The second plot shows the number of genes phased by a single phase-block across different phasing percentage categories (0%, 0-25%, 25-50%, 50-75%, 75-100%, and 100%) on the x-axis, with the y-axis showing the count of genes per individual within each phasing category. **B)** In proband DSDTRN17, LRS resolved pathogenic compound heterozygous variants in the Luteinizing hormone/choriogonadotropin receptor (*LHCGR*) gene, causing Leydig cell hypoplasia. The first figure illustrates the variants’ locations on the protein; Exon 9 deletion truncates the extracellular domain, while the exon 11 missense variant is situated in a transmembrane region with other known pathogenic variants.

Using our clinical annotation and prioritization pipeline on these long-range phased variants, we identified clinically relevant compound heterozygous variants in four probands (1 diagnostic and 3 candidates requiring additional evidence). In proband DSDTRN17, we found compound heterozygous pathogenic variants in the Luteinizing hormone/choriogonadotropin receptor (*LHCGR*) gene associated with recessive Leydig cell hypoplasia **(Figure 6B)**. The first variant was a pathogenic/likely-pathogenic missense SNV located on exon 11, annotated in ClinVar (p.Ser616Tyr, chr2:48915089G>T, Variation ID: 14393) and identified by SRS. The second variant was an LRS-resolved 6,694 bp deletion, which deleted exon 9 of the gene. Through LRS phasing, we successfully identified these compound heterozygous variants in the absence of parental DNA, providing a definitive diagnosis.

### Haplotype-specific methylation profiling with LRS can prioritize rare intronic SVs and variants of uncertain significance (VUS)

Single-molecule LRS captures both DNA sequences and modifications simultaneously (Logsdon, Vollger, and Eichler 2020). Napu runs modkit (https://github.com/nanoporetech/modkit), which generates haplotype-specific and combined CpG methylation calls. We used these calls to identify methylation outlier regulatory regions in probands, potentially linked to rare, but previously unprioritized variants in a haplotype-specific context. We analyzed regional methylation (methods) in CpG Islands (CGIs) and ENCODE cis-regulatory elements (cCREs) across individual haplotypes. To account for variability in large CGIs, CpGs were segmented by methylation patterns, with segment consistency maintained across samples (methods).

Differentially methylated CGIs (DM-CGIs) were identified per proband haplotype using all unaffected, unrelated parents (control set 1) and other probands (control set 2) as controls (methods). A median of 11 DM-CGIs per proband were found, including 2 near (± 10 Kbp) disease genes, 3 near other protein-coding genes, and 5 without any protein-coding gene nearby **(Figure 7A)**. Similarly, out of a median of 22.5 DM- CCREs called, 10 were near (± 1 Mbp) disease genes, 11 near other protein-coding genes, and 1 without any protein-coding gene nearby (**Supplementary Figure 18**). For a subset of these differentially methylated regions (DMRs) within ≤10kb of genes expressed in blood, we found a significant association between methylation and gene expression changes (**Supplementary Figure 19)**.

**Figure 7:**
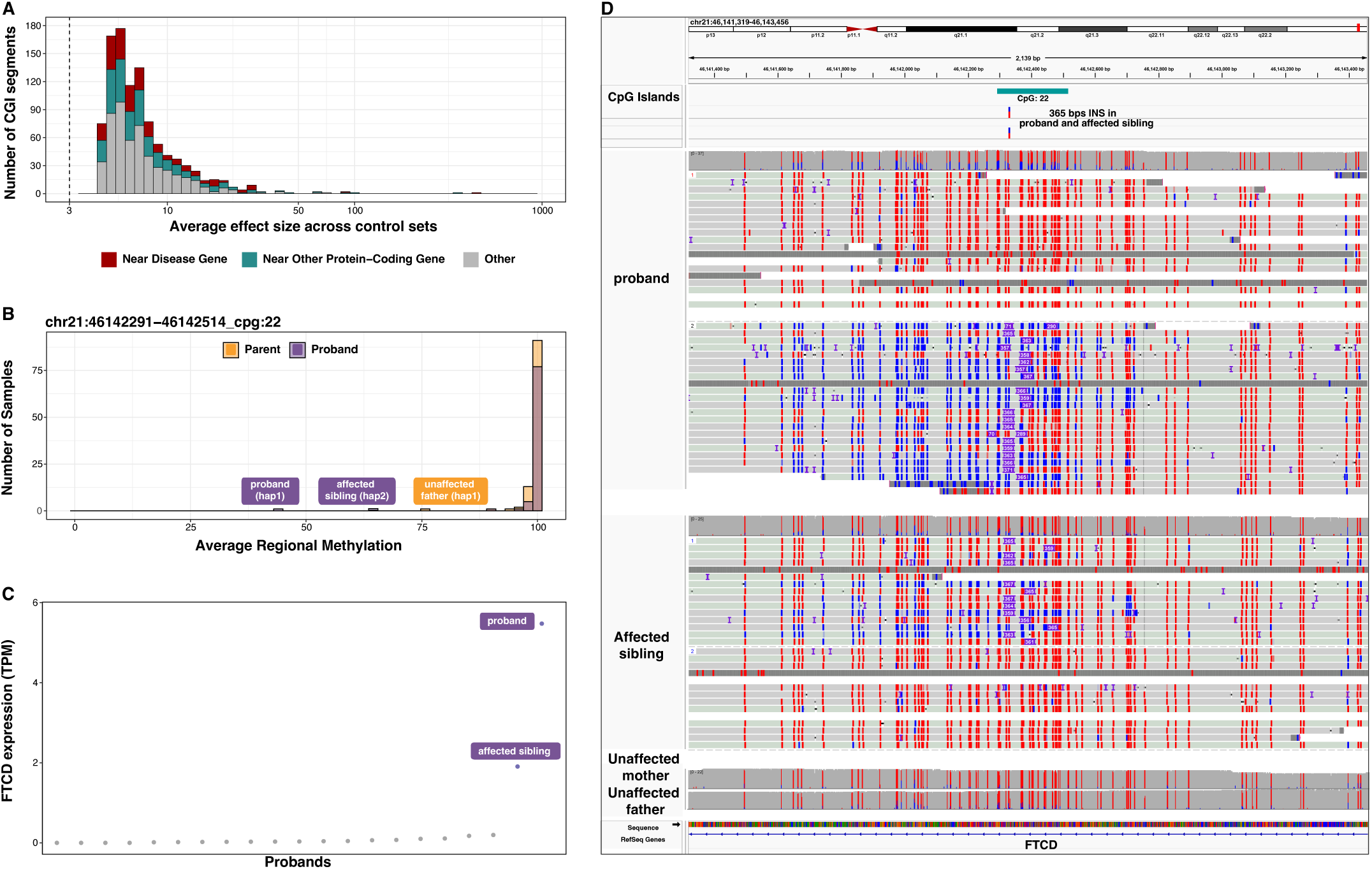
Haplotype-specific methylation profiling with LRS. **A)** Haplotype-specific DM-CGIs distributed by average Cohen’s d across control sets in all probands. The vertical dotted line highlights average Cohen’s d cutoff for significant DM-CGIs per proband haplotype, computed from the methylation profile of controls (control set 1) and remaining probands (control set 2). CGIs near (within ± 1 Kbp) known disease genes are highlighted in red, near other protein-coding genes in teal and others in grey. **B)** Average methylation at DM-CGI in *FTCD* across all samples, shows hypomethylation in both affected siblings. The unaffected father shows slightly higher methylation. **C)** *FTCD* expression TPM values across 21 affected individuals with RNA-Seq data from blood shows overexpression in both affected siblings. **D)** IGV shows methylation status of CpG sites in DM-CGI across both affected siblings and a 365 bps insertion (5 copy expansion of a VNTR) likely causing hypomethylation of CGI in *FTCD*. Blue indicates unmethylated CpG prediction and red indicates methylated CpG prediction with color intensity corresponding to base modification probabilities.

A clinical review of these DMRs did not allow us to directly implicate any in a participant’s phenotype, however it did identify interesting correlations. For example, in a case involving two affected siblings, we identified a haplotype-specific hypomethylated CGI in the *FTCD* gene **(Figure 7B)**. Although *FTCD* is typically expressed at very low levels in blood, it was found to be overexpressed in both siblings, correlating with hypomethylation **(Figure 7C)**. The hypomethylation and corresponding overexpression is likely caused by a 365 bps rare intronic LRS-only SV insertion at the same position in the DM-CGI **(Figure 7D)**. This region is a variable number tandem repeat (VNTR) with a repeat unit size of 73 bp, exhibiting a 5 copy expansion in the affected siblings. Shorter insertions of 146 bp (2 additional copies) were detected at the same position in 11 unrelated samples with a normal degree of methylation and expression. The unaffected father also carried the insertion (albeit with a slightly higher methylation signal), but no RNA-seq data was available to assess expression impacts in the father. While the variant is unlikely to be contributing to the phenotype, this family highlights that methylation alterations can identify regions with DNA variants and altered expression that should be further studied.

### Validation of causal variant with an independent evaluation of known episignatures using ONT methylation

In one case, PMGRC-146-146-0 (a proband with a complex neurodevelopmental phenotype), analysis from SRS identified a *de novo* deep intronic single nucleotide variant of uncertain significance in *ARID1B*, ARID1B:c.3025+700C>G. This variant had moderate predictions by Splice AI of leading to an acceptor gain (Splice AI: AG 0.31). Using the LRS data, we confirmed this *de novo* finding and interrogated the percent methylation at 82 CpGs, previously reported by Aref-Eshghi et al in 2018 (Aref-Eshghi et al. 2018) as being differentially methylated in Coffin-Sirus Syndrome 1 (CSS1). Through permutation testing, we demonstrated that the methylation pattern for this sample specifically matched the CSS1 episignature and was distinct from the other samples in our set. Integrating this molecular phenotype with the molecular data and transcriptomic data confirmed the diagnosis and permitted the reclassification of the variant to Likely Pathogenic under current ACMG variant classification guidelines.

### Summary of Diagnostic Findings

Out of 42 affected individuals in this cohort, diagnostic variants were found for 11 of the probands, (**Supplementary Table 24**). As reported in detail in earlier sections, these include 4 probands with *de novo* variants (one validated by a matching episignature), 1 proband with compound heterozygous variants, 2 probands with sex-chromosomal translocations and 4 probands with complex compound heterozygous variants (including gene fusions) in a complex segmental duplication containing *CYP21A2.* Additional candidates that require further evidence were identified for a further 4 probands (1 *de novo* variant, 3 compound heterozygous variants) (**Supplementary Table 25**). The remaining 27 affected individuals remained unsolved with ongoing strong suspicion for a monogenic cause.

Our cohort consisted of two distinct subsets of affected individuals. To test the ability of the method to detect known variants of clinical significance, the first set consisted of 6 individuals with complex SVs, including known translocations or variants in segmental duplications, which were either detected with targeted clinical tests or weren’t previously fully resolved at the base-level. In all of these cases, LRS was able to characterize the causal variant (2 with sex-chromosomal translocations and 4 with complex compound heterozygous variants in *CYP21A2-CYP21A1P*) precisely. The second set consisted of 27 individuals with a high clinical suspicion of monogenic disease and no prespecified genetic locus of interest for inspection by LRS. These included mainly but not exclusively individuals with neurodevelopmental phenotypes, specifically selected for inclusion to enrich for likely *de novo* causal variants (McRae et al. 2017) (**Supplementary Table 26-27**). The final set included 9 individuals with inconclusive SRS findings **(Supplementary Table 28)**. By genome-wide analysis of rare (inherited and *de novo*) small variants, SVs, TREs, and methylation outliers, causal *de novo* variants were identified in a total of 4 probands, 3 of which were also detectable by reanalysis of the SRS data (*RNU4-2* in two probands, *GLUL* in one proband) and 1 also detected by SRS yet only validated by confirming a known epigenetic signature for *ARID1B* accessible by LRS-only. Biallelic variants in *LHCGR* were resolved in 1 case using LRS-only. In addition, candidates were identified in a further 4 probands (*BLOC1S1*, *SLC6A3*, *SRSF2,* and *LHCGR*), of which in 2, the variant of interest was detectable by LRS-only (55 bp intronic deletion in *SLC6A3* and 300 bp upstream insertion in *LHCGR*).

## Discussion

In this study, we sequenced an undiagnosed rare disease cohort using a time and cost efficient, one flow cell nanopore sequencing protocol, yielding on average ∼113 Gb of ONT reads (corresponding to ∼36x coverage). The Napu pipeline was used to process the sequencing data to generate assemblies, variants, phasing, and methylation calls in a single run. Both our wet and computational protocols are openly available (methods), with the latter being reproducible on both cloud and local infrastructure quickly and inexpensively (Schatz et al. 2022).

We evaluated the additional yield of LRS by systematically comparing it with SRS. Our results reinforce earlier findings that LRS can address more of the genome (Aganezov et al. 2022; Kolmogorov et al. 2023) and broadly demonstrate that this translates into additional clinically relevant information. LRS accurately mapped many genomic regions without coverage in SRS, providing resolution in coding exons of a median of 280 protein-coding genes per individual missed by SRS. Some of these genes overlapped highly identical segmental duplications, known to be implicated in Mendelian disorders, but which have been refractory to SRS detection (Wagner et al. 2022; Trier et al. 2020; Awdeh and Alper 1980; X. Chen et al. 2020). Indeed, most LRS-exclusive SN-DNVs overlapped low complexity and low short-read mappable regions. Also supporting the idea that LRS provides additional clinical value, despite higher base-level error rates, we find that LRS identifies many additional rare functionally annotated small variants relative to SRS. Interestingly, the additional number of such variants relative to SRS increased substantially when reducing the threshold for genotype quality filtering, and simultaneously, the number of SRS-exclusive functionally annotated variants decreased. While reducing the quality filter will likely introduce some false positives, it is likely that further technology developments to improve the base-level accuracy of ONT long reads will result in still larger gains in the number of confidently ascertained functionally annotated variants relative to short reads. Finally, and also reinforcing many earlier studies, we find huge numbers of additional SVs relative to SRS, including rare tandem repeat expansions in protein-coding genes (Jensen et al. 2024; Medhat Mahmoud et al. 2019). The quality of these SV calls is clearly superior to what has been possible with SRS. Overall, our comparisons reinforce the strengths of LRS in accurate mapping to new and previously inaccessible regions, with biggest gains evident in accurate, base-level SV characterization.

Our clinical diagnostic results show the value of both the discovery of LRS-exclusive candidates and the re-prioritization of candidates found by both SRS and LRS, by virtue of reanalysis. Consistent with prior reports, reanalysis has considerable value (Wijngaard et al. 2024; Schobers et al. 2022; van Slobbe et al. 2024), as novel pathogenic variants, disease-associated genes, and additional disease mechanisms or inheritance modes are continuously being discovered (as seen for *GLUL* in one of our cases). We suspect that many of the remaining undiagnosed cases in our cohort are also caused by highly penetrant monogenic variation that we have not yet been able to confidently interpret and can hopefully be addressed with future re-analyses.

Related to the ongoing value of reanalysis, we found many variants with LRS that we could not usefully interpret. Clearly, to more fully utilize the additional information provided by LRS, more comprehensive population and clinical databases based on long-read data are needed. Currently large reference population databases are short-read based and hence lack population-based allele frequencies for LRS-derived variants (Collins et al. 2020; Karczewski et al. 2020). Similarly, clinical databases like ClinVar (Landrum et al. 2014) contain variants that have almost exclusively been detected by short reads (plus other clinical genetic tests) and do not yet contain the classifications of many "long-read exclusive’’ variants to help with their identification and prioritization. Due to this, most of the LRS-exclusive rare SVs, tandem repeat expansions, and small variant FAVs are currently difficult to interpret. This somewhat frustrating situation will only be solved by the creation of LRS-derived databases of variants. There is a need for diverse, population-scale sequencing of healthy cohorts to create repositories of variation and their frequencies. Projects like the Human Pangenome Reference Consortium (Liao et al. 2023), the Human Genome Structural Variant Consortium (Ebler et al. 2022), and larger biobank scale projects, like those from the All of Us project (M. Mahmoud et al. 2024) are starting to generate the much-needed sequencing data for this. In tandem, and likely over a much longer timescale, the expansion of LRS in clinical genome sequencing should progressively lead to the discovery and contribution of disease-causing variants to clinical databases.

LRS facilitated the prioritization of rare, non-coding SVs by providing complementary methylation information and enabled the detection of methylation outliers, some of which correlated with gene expression, though none were definitively diagnostic in this cohort. Methylation information provided through LRS also provided orthogonal validation using known episignatures in one of our cases (Li et al. 2023; Martin-Trujillo et al. 2020; Chundru et al. 2023). Interpreting rare methylation outliers is challenging due to the complex interplay of environmental and genetic factors, and the lack of good reference datasets. However, more LRS- based clinical studies analyzing methylation in conjunction with genetic variation in rare diseases will help establish the increased methylation-based diagnostic yield (Cheung et al. 2023).

Our *de novo* comparison revealed a potential application of LRS for distinguishing postzygotic mosaic variants from prezygotic *de novos*, which may in the future contribute further understanding to rare genetic diseases (Tinker et al. 2023; Noyes et al. 2022) and the penetrance (or lack of) for such variants. Related to this, our future work will focus on developing accurate characterization methods for comparing *de novo* indels between sequencing technologies to avoid discrepancies and prevent missed or falsely identified variants.

Overall, LRS enabled more comprehensive genome analysis. Decreasing costs of LRS, leveraging cost- efficient sequencing and computational protocols, coupled with clinical analysis pipelines for analyzing all alteration types together, should lead to accurate diagnoses for individuals with suspected Mendelian conditions who today remain unsolved after a comprehensive evaluation. To more precisely estimate the benefits of whole genome LRS relative to SRS will require larger cohorts, nuance in appreciating the phenotypic makeup of the cohort, and will likely change over time with continued data sharing to progressively overcome the interpretation gaps that this new data reveals.

## Methods

### Overview of sequencing protocol

Isolated DNA, white blood cells, or whole blood samples were received from Broad Institute and Children’s National Hospital (CNH). CNH samples were either from the DSD-Translational Research Network biobank or from the PMGRC/UCI site of the GREGoR Consortium. Some had high molecular weight (HMW) DNA previously extracted for Optical Genome Mapping on the same samples, using the Bionano recommended protocol. Twenty probands with neurodevelopmental phenotypes, one affected sibling, and forty unaffected parents (n=61) were consented to the Broad Institute Rare Genomes Project (RGP), which includes the use and sharing of data for research purposes (Mass General Brigham IRB protocol 2016P001422) and had previous srWGS sequencing and analysis performed. Two EDTA tubes (4 mL) and one PAXgene RNA tube of whole blood were collected from participants from across the United States and sent at room temperature by overnight courier to the Broad Institute or CNH. Upon receipt, samples were frozen, and an EDTA tube from each participant was later shipped in bulk on dry ice to UCSC.

Of the 105 samples received at UCSC, 6 samples with pre-extracted DNA were excluded from the cohort for not meeting our quality standards for HMW DNA size. Early in protocol development, five whole blood samples failed DNA extraction due to low yield. Four of these were replaced and successfully sequenced, resulting in a total of 98 sequenced samples. HMW DNA was extracted using Circulomics Nanobind CBB Big DNA Kit (NB-900-001-01) or NEB Monarch HMW DNA extraction kit for cells and blood (NEB T3050). Approximately 5 µg of isolated DNA was sheared using Diagenode Megaruptor 3, DNA fluid+ kit (E07020001). The size of sheared DNA fragments was analyzed on the Agilent Femto Pulse System using genomic DNA 165 kb kit (FP-1002-0275). Fragment size distribution of post-sheared DNA had peak at approximately 50kb. Small DNA fragments were removed from the sample using PacBio SRE (Short Read Eliminator) kit (SKU 102-208-300). Library preparation was carried out using Oxford Nanopore Technologies (ONT) ligation sequencing kit V14 (SQK-LSK114). Sequencing was performed on the PromethION 48 sequencer using R10.4.1 flow cells. Each sample was used to prepare four libraries per flow cell. Flow cells were washed using the ONT wash kit (EXP-WSH004) and reloaded with a fresh library every 24 hours for a total sequencing runtime of 96 hours. Human bulk transcriptome sequencing was performed by the Genomics Platform at the Broad Institute of MIT and Harvard. The transcriptome product combines poly(A)-selection of mRNA transcripts with a strand-specific cDNA library preparation, with 150 bp reads and a mean insert size of 550 bp. Libraries were sequenced on the HiSeq 2500 platform to a minimum depth of 75 million STAR-aligned reads.

We then ran the Napu end-to-end pipeline on 98 samples, generating diploid *de novo* phased assemblies, harmonized variant calls against the GRCh38 reference genome (merging reference-based small variant calls and assembly-based SV calls), and haplotype-specific methylation calls.

### Genome Completeness Analysis

To identify genomic regions uniquely callable by LRS and SRS, we used BAM files aligned to GRCh38 and T2T-CHM13v2.0 reference genome. Assembly gaps and simulated centromeric regions in the GRCh38 reference were excluded from the analysis to avoid false coverage estimates. Since raw data for SRS wasn’t available to us, we reverted GRCh38-aligned SRS CRAMs and remapped them to T2T-CHM13 using Picard RevertSam (v1.141). We then analyzed per-base depth using mosdepth (v0.3.4). Aligned reads with a minimum mapping quality of 10 were considered (90% probability of correct mapping), and coverage bins were defined as 0:1:10:80, using the ′*--quantize*′ option to merge adjacent bases within the same coverage bins. Bases with depth 0 were binned as NO_COVERAGE, 1-9 as LOW_COVERAGE, 10-79 as CALLABLE, and >=80 as HIGH_COVERAGE. The Gencode Comprehensive gene set release 45 for GRCh38 was used to define protein-coding genes (includes exons and introns) and coding exons. Mendelian disease genes were genes in OMIM (Hamosh et al. 2005) and known disease genes from ClinGen (Rehm et al. 2015) and GenCC (DiStefano et al. 2022). Ideograms were plotted using KaryotypeR (Gel and Serra 2017).

### SV-aware SnpEff-based Clinical Annotation Workflow

We developed a clinical annotation workflow to annotate SNVs, indels, and structural variants from Napu harmonized VCF. After splitting multi-allelic variants with bcftools (Danecek et al. 2021), they were annotated with SnpEff (Cingolani et al. 2012) v5.1 using the GRCh38.105 pre-built database. Small variants (30 bp or less) and structural variants were then annotated separately.

Structural variants were annotated using the *sveval* package (Hickey et al. 2020) to add frequency estimates and flag SVs that overlapped with known clinical SVs (nstd102 in dbVar) or any DGV SVs (GRCh38_hg38_variants_2020-02-25.txt). To annotate their frequency, each SV was matched with SVs in eight public SV databases (DGV common SVs/nstd186, gnomAD-SV/nstd166, Sirén et al. 2021, HPRC v1.0, HGSVC2, 1000GP ONT Miller, 1000GP ONT Vienna, 1000GP ONT SV imputation panel), if their reciprocal overlap was at least 50% and they were located at less than 100 bp. Of note, the simple repeat track from the UCSC Genome Browser was used to add a wiggle room to help match SVs placed differently around repeats. SVs were considered rare if the frequency of all matched variants in all databases was below 1%. SVs were also annotated with the number of enhancers of known disease genes that they overlapped, using GeneHancer and ENCODE ELS lists as enhancer catalogs. In parallel, AnnotSV (Geoffroy et al. 2018) was used to annotate SVs using the GRCh38 human pre-built database and GeneHancer (v5.9, (Fishilevich et al. 2017)). When HPO terms were available, they were provided to AnnotSV. Finally, SVs were re-genotyped from the raw long reads using vg (Hickey et al. 2020) to count the number and proportion of reads supporting the alternate allele. SV calls with no supporting reads were filtered out.

After the first SnpEff annotation, small variants were further annotated with SnpSift v5.1 to flag ClinVar variants (2023-03-18 version), annotate their frequency in gnomAD v3.0, as well as GERP++, CADD, MetaRNN, and ALFA provided through dbNSFP (v4.4, (Liu et al. 2020)). SnpSift was also used to keep only small variants with "HIGH" or "MODERATE" impact, or a percent of loss-of-function transcript higher than 0.9, or those that are in ClinVar.

For variant curation, we also integrated gene-level information such as pLI scores from gnomAD v2.1.1, the dosage sensitivity map from (Collins et al. 2020), genes in OMIM, and known disease genes from ClinGen and GenCC. The workflow described above is available in Snakemake and WDL formats at https://github.com/jmonlong/variant_annotation_wf.

### Family-based analysis of LRS in *seqr*

SNVs and indels called by DeepVariant for the 20 RGP families (19 trios, 1 quad) were loaded to the *seqr* genomic analysis platform for family-based monogenic disease analysis (seqr.broadinstitute.org/). Briefly, “*De Novo*/Dominant” and “Recessive” variant searches with both “Restrictive” and “Permissive” thresholds for reports of pathogenicity, functional consequence and predicted deleteriousness, allele frequency (in the gnomAD reference population (S. Chen et al. 2024)), and call quality we applied as reported in detail by Pais et al (Pais et al. 2022). The returned variants were assessed for relevance to reported phenotype and disease mechanism (e.g., loss-of-function, gain-of-function) using the ClinVar (Landrum et al. 2014), OMIM (Amberger et al. 2015), and DECIPHER (Firth et al. 2009) databases, in addition to the potential to cause novel disease-associations using external resources linked to *seqr*, spanning gene-level data (e.g., gnomAD constraint metrics (Karczewski et al. 2020)), tissue expression data from the Genotype-tissue Expression (GTEx) Portal (Lonsdale et al. 2013), and functional data (e.g., mouse models), among others.

### *De novo* pipeline for LRS small variants

To identify putative *de novo* small variants (SNVs and indels < 30 bps) from LRS, individual harmonized VCFs from Napu for a trio were preprocessed to retain PASS calls and non-homozygous reference variants. This was followed by running rtgtools vcfeval (v3.12.1) in a pairwise manner, to identify variants that were called in the child but not in either parent. Variants with GQ<20, DP<10, or homozygous alternate calls were removed. For male samples, homozygous alternate variants in sex chromosomes with DP>=5 were retained. Next, rare variants (AF<0.001 as per gnomAD v3.0) were selected. These constituted the “whole-genome” callset. The “annotated” callset included non-intergenic variants, predicted by SnPEff to have some impact on overlapping or nearby genes. More annotations including GERP++, CADD, and MetaRNN provided through dbNSFP (v4.4) were added. Further, to generate a stringent set of DNVs, we utilized an in-house script for in-silico read validation using parental reads. Only variants with no read-support in both parents were retained. Finally, DNVs called in more than one individual from the entire cohort were removed. The workflow described is available in a WDL format at https://github.com/shlokanegi/denovo_smallvars.

### *De novo* pipeline for SRS small variants

SRS was performed on DNA purified from blood by the Broad Institute Genomics Platform on an Illumina sequencer to 30x average depth. Raw sequence reads were aligned to the GRCh38 reference genome. Variants were called with GATK version 4.1.8.07 in the form of SNVs and indels < 30 bp to generate a joint called VCF file. To identify putative *de novo* variants, the VCF file was loaded with Hail (https://github.com/hail-is/hail), and Hail’s de_novo() function was called with variant allele frequencies from gnomAD GRCh38 exomes v2 and genomes v3 used as priors. After calling, putative *de novo* variants were excluded from the dataset if present in gnomAD or within their own call set at an allele frequency ≥0.1%, containing a GATK Variant Quality Score Recalibration (VQSR) flag in the filters field of the VCF, falling within a reported problematic region of the genome (downloadable from the UCSC browser: ucscUnusualRegions.bed, encBlacklist.bed, grcExclusions.bed), falling within close proximity to other *de novo* variants in the same individual (within 1 Kb), or having a call rate of ≤0.99 in the call set. Variants were also excluded if the proband had <5 alternative or <5 reference reads or an allele balance <0.2 (<0.3 for indels), or if any of the proband, mother, or father had a depth <10 (<15 for indels) or a genotype quality <20 (<25 for indels).

### Validation categories for SRS and LRS DN-SNVs

To assess efficacy of LRS in detecting additional rare DNVs as compared to SRS, we applied the LRS *de novo* calling pipeline for small variants to 21 trios from 20 families (19 trios and 1 quad, using both affected siblings in the quad as separate trios) with available data. SRS rare DNVs were generated with a separate SRS *de novo* pipeline. For ease and strictness of comparison, analysis was restricted to SNVs. These were filtered out if called in >1 family or showed evidence in even a single read in parental samples (a high level of stringency, but potentially excluding cases of parental mosaicism and those also including recurrent sequencing errors).

Categorization criteria for defining validation category for technology-specific exclusive DN-SNVs are; 1 - Called by other technology as a non-DNV - “Called by other technology’s variant caller, but classified as non- DNV”. They were either present in one/both parents or were not unique within the cohort. 2 - “Likely False-positive” - False-positive due to multiple reasons. AB<0.2, multi-allelic, region noisy with multiple indels on affected haplotype, or low-quality bases. 3 - “Likely post-zygotic mosaic” - Discordant DN-SNVs (SRS-only DN-SNVs excluding category 1 and 2) were classified as likely postzygotic mosaic, if a) LRS phasing showed the presence of at least three reference alleles on the alt-haplotype, b) Phasing was accurate, confirmed by at least two surrounding heterozygous SNVs on both sides c) Absence of a third allele when mapped to T2T- CHM13 or GRCh38 d) Absence of INDELs on reads phased to alternate haplotype, which could be a homopolymer or short tandem-repeat errors on long reads, and e) AB on both SRS and LRS was < 0.5. 4 - “Likely true-positive” - All the remaining exclusive DN-SNVs with AB>=0.2.

### *De novo* pipeline for LRS Structural Variants

A first list of *de novo* structural variant candidates was produced by comparing the SV LRS calls in the proband and their parents using the *sveval* package. As above, SVs are matched based on their location, type, and size, with some added wiggle room in annotated simple repeats (UCSC Genome browser track). Looser matching thresholds (5% reciprocal overlap, 500 bp distance) were used to produce a stringent list of *de novo* candidates by ensuring that no similar SVs were called in the parents. These *de novo* SV candidates were then re-genotyped in the proband and the parents using vg and the raw long reads. We kept variants that were supported by at least 25% of the reads in the proband, and less than 10% in both parents. Of note, variant calls overlapping an assembly gap in GRCh38 (gap track from UCSC Genome Browser) were removed.

### Short tandem repeat expansion

Phased SV calls by Hapdiff were clustered based on the simple repeat annotation from the UCSC Genome Browser (simpleRepeat track). For each annotated site, we retrieved how many bases were deleted or inserted within the repeat region, separately for each haplotype.

We ran the same analysis on two public datasets of healthy individuals to use as controls when looking for outliers (see below). First, phased SVs derived from the Human Pangenome Reference Consortium (HPRC) v1.0 pangenome made with Minigraph-Cactus were extracted from hprc-v1.0-mc.grch38.vcfbub.a100k.wave.vcf.gz VCF file, using only variants larger than 30 bp. Second, phased SVs derived from the Human Genome Structural Variant Consortium (HGSVC) assemblies, freeze 3, were extracted from the hgsvc.freeze3.sv.alt.vcf.gz VCF file.

For each simple repeat site, we compared the size delta (number of bases deleted/added) in each proband, to the full cohort and the HPRC/HGSVC controls. We computed a Z-score for each as the value for the proband minus the average across controls, divided by the standard deviation across the controls. We then selected the least extreme Z-scores of the two. Hence, a Z-score higher than 5 means that the site is expanded above 5 standard deviations compared to both the cohort and the HPRC/HGSVC controls.

Each region was annotated with the distance to the nearest coding exon defined in Gencode V41. We annotated the nearest protein-coding gene and the nearest gene known to be associated with a disease.

### Identification of sex chromosomal translocations

The Napu pipeline produced reads aligned to the GRCh38 reference genome. We first ran mosdepth (v0.3.4) on these BAM to identify abnormal copy-number in two probands. Specifically, we observed coverage suggesting two copies for most of the chromosome X but also evidence for one copy of a small part of chromosome Y, including the *SRY* gene. For those two samples with suspected translocations, we re-mapped the reads to the T2T-CHM13 genome and found evidence of the translocation in the form of long split-mapped reads where each part mapped on each side of the approximate breakpoints suggested by the read coverage analysis. Those supporting reads were collected using an in-house script that identified the reads alignments breaking around the location of copy number changes. Thanks to the split-mapped reads, both breakpoints of the translocation were located at base-level resolution. Of note, we also tested calling the translocation using Sniffles2 on reads mapped to the T2T-CHM13 genome using minimap2 or NGMLR.

### Detecting gene fusion and conversions of CYP21A2

We developped a pangenome-based approach, called Parakit to characterize the complex locus hosting the CYP21A2 gene (manuscript in preparation). A local pangenome is first built, collapsing the module with the pseudogene and the module with *CYP21A2* in the GRCh38 reference. High quality assemblies produced by the Human Pangenome Reference Consortium are also integrated in this pangenome which is then used as a reference to re-align reads from this region. Because the similar copies are collapsed in the pangenome, the reads now align to only one position. Furthermore, they traverse the pangenome through nodes that are informative for inference because they are specific to the CYP21AP pseudogene or CYP21A2 gene modules. Using this information, Parakit analyzes the read alignments on the pangenome graph to identify and visualize reads supporting fusion/gene conversion events and the corresponding changes in read coverage or allelic balance. Parakit also infers the most likely haplotypes by enumerating walks through the pangenome graph that are consistent with the aligned reads. Parakit will be available on GitHub soon (manuscript in preparation).

### Annotation, segmentation, and average regional methylation calculation within CpG islands and cis-regulatory elements

Within Napu, modkit was run with the default --filter-threshold of 10 percentile, with the --cpg, --collapse- strands, and --partition-by HP parameters. The generated haplotype-resolved bedMethyl files were used to calculate regional methylation in pre-defined regions of interest. We used CpG Islands (available from the UCSC browser) and human cis-regulatory elements from ENCODE Registry of CCREs V3 as target regions. Large target regions can often represent variable methylation which will be incorrectly captured by regional average methylation calculation. To tackle that, we developed SegMeth (https://github.com/shlokanegi/SegMeth), which segments target regions into windows that exhibit statistically different methylation patterns. SegMeth was run with relaxed parameters to avoid over- segmentation (-p 1e-7, -ut 30, -mt 70, -minCG 5). On average, over 1,300 CGIs exhibited variable methylation and could be segmented into two or more statistically significant segments **(Supplementary Figure 20)**. Segments were generated for all 98 samples, and collapsed to generate intersected segments using an in-house script. Gencode’s comprehensive gene set, release 45, was used to annotate all target regions (CGI segments and CCREs) using bedtools closest and bedtools groupby (v2.29.1). Each target region was annotated with the nearest coding exon(s) and gene(s) (within ±1 Mbp for CCREs and ±1 Kbp for CGIs). A higher distance threshold for CCREs was considered due to the presence of enhancers in the database, some of which are known to exert distal effects on target genes by DNA looping in 3D chromatin space (Pennacchio et al. 2013).

Average methylation across annotated target regions was calculated for all samples using both haplotype- specific bedMethyl pileups. For each target region, an average of %methylation was taken across all individual CpGs with atleast 5x valid coverage. Regions in samples with <10 CpGs or >50% of low-coverage CpGs were flagged to aid with downstream processing and DMR calculations. The workflow described above is available as a WDL at https://github.com/nanoporegenomics/Napu_wf/blob/r10/wdl/workflows/bedtoolsMap.wdl.

### Detection of differentially methylated regions

For each regional target (CGI segments and CCREs), the average methylation values of proband haplotypes were systematically compared to those of two control sets (control set 1: all healthy parents; control set 2: remaining probands) using paired t-tests. Multiple testing corrections were applied using the Benjamini- Hochberg (BH) method. Cohen’s d was reported as a measure of effect size, and the methylation difference between the proband and the control means was used to filter out statistically significant but biologically irrelevant DMRs. Only target regions with regional methylation measurements in at least half the samples in the cohort were tested. Significant DMRs were identified using these criteria: effect size ≥ 3, BH-corrected p- values < 0.001, and mean methylation difference ≥ 40% for both proband vs. control set 1 and proband vs. control set 2 comparisons. To identify potential disease-associated DMRs per proband, we inferred their transmission status (*de novo* across all samples, or inherited from one or both parents). This allowed for further investigation into rare variants within or near DMRs, in context of haplotype-specificity as well as transmission pattern. A DMR with "*de novo* transmission" had one or both haplotypes clear outliers (hyper or hypo) when compared to all other samples. A DMR with “inherited transmission” had one of both parents clear outliers along with proband.

## Data Availability

All data produced in the present study are available upon reasonable request to the authors

## Acknowledgements

The Chan Zuckerberg Initiative (CZI) provided funding for the sample collection, sequencing and analyses described. We would like to especially acknowledge the support of Dr. Jonah Cool, Dr. Sara Simmonds, and Mr. Bruce Martin who provided helpful feedback at various points throughout this project. We thank Dr. Paolo Carnevali for his assistance and support on the use of the Shasta genome assembler. BP and SN were also supported by National Institutes of Health (NIH) grants R01HG010485, U41HG010972, U24HG011853 and OT2OD033761. M.K. was supported in part by the Intramural Research Program of the NIH. AODL and RGP SRS and analysis were supported in part by NIH grants UM1HG008900, U01HG011755 and R01HG009141 and grants 2019-19927, 2020-224274, 2022-316726, DOI https://doi.org/10.37921/236582yuakxy, from the CZI DAF, an advised fund of Silicon Valley Community Foundation (funder DOI 10.13039/100014989), and in part by research funding from Illumina Inc. SLS was supported by a fellowship from the Manton Center for Orphan Disease Research at Boston Children’s Hospital and GL by a fellowship from Fonds de recherche en santé du Québec (FRQS). Samples were collected from the DSD-Translational Research Biobank (supported by R01HD068138 and RO1HD093450; EV, ED) and the GREGoR-UCI/CNH site (supported by U01 HG011745, EV, ED, SB). For the DSD-TRN samples, WGS was funded in part by the Gabriella Miller Kids First Initiative grant X01HL132384. We gratefully acknowledge the help of DSD-TRN investigators Meilan Rutter, Phyllis Speiser, Natalie Nokoff, Courtney Finlayson, Jodie Johnson, who contributed the diagnosed samples used here and of Miguel Almalvez (UC Irvine) for management of the DSD-TRN and UCI-GREGoR biobanks and sample extraction. All sample, family, and individual IDs used in this study have been fully de-identified and are not known to anyone outside the research group.

## Author contributions

B.P., J.M., K.M., A.D. and E.D. helped conceive and direct the study. S.N. and J.M. performed data analysis. S.S. and E.C. contributed to RGP cohort analysis and S.B. contributed to PMGRC sample clinical analysis. B.M., J.G., T.H., S.R. contributed to ONT data sequencing. I.V. contributed to basecalling. M.L., G.V., S.J., B.M., and G.L. were involved in recruitment and selection of RGP clinical cases for sequencing. S.N., B.P., J.M. and S.S. drafted the manuscript.

## Supplementary Figures

**Supplementary Figure 1:**
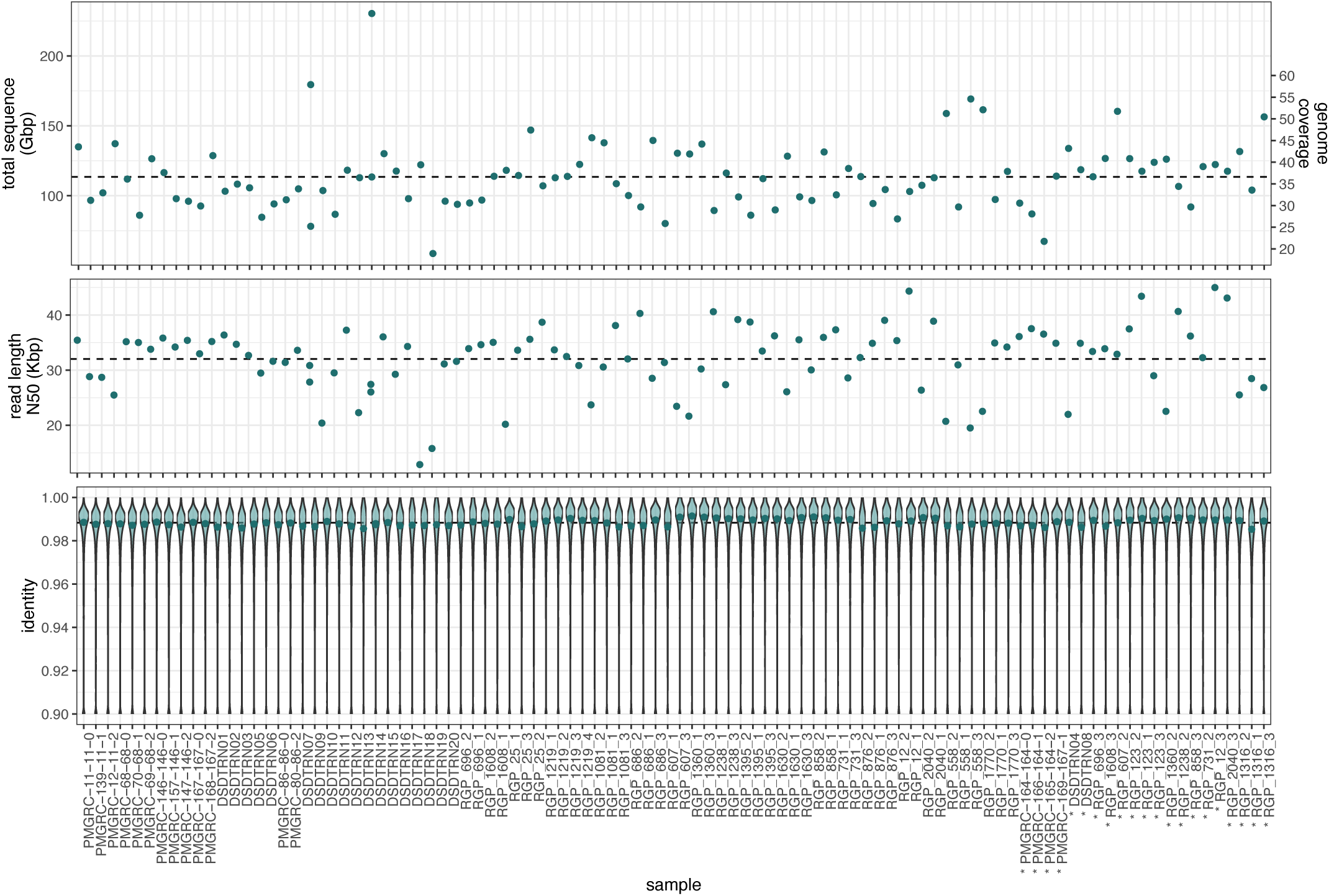
Sequencing results per sample. Samples sequenced with 2 flow-cells are denoted with an asterisk (*).

**Supplementary Figure 2:**
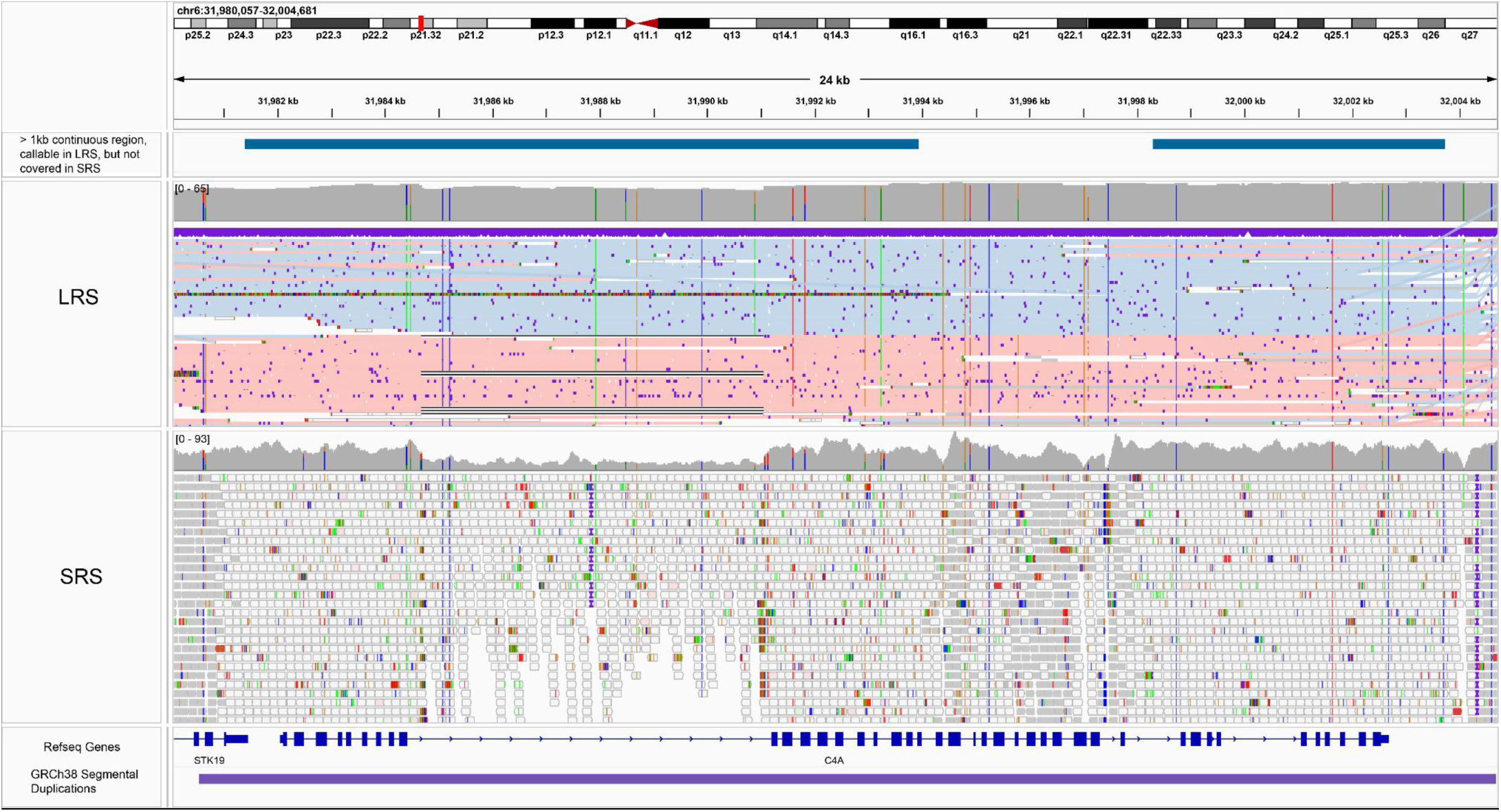
C4A gene length exclusively callable by LRS. The top track shows >1 kb continuous region which was callable in LRS, but had no coverage in SRS. In LRS track, reads are sorted and colored by haplotype. In SRS track, white transparent reads have 0 mapping quality, i.e. are not uniquely mapping to the region. The bottom track (purple) is the segmental duplication annotation from GRCh38 GIAB stratification callset.

**Supplementary Figure 3:**
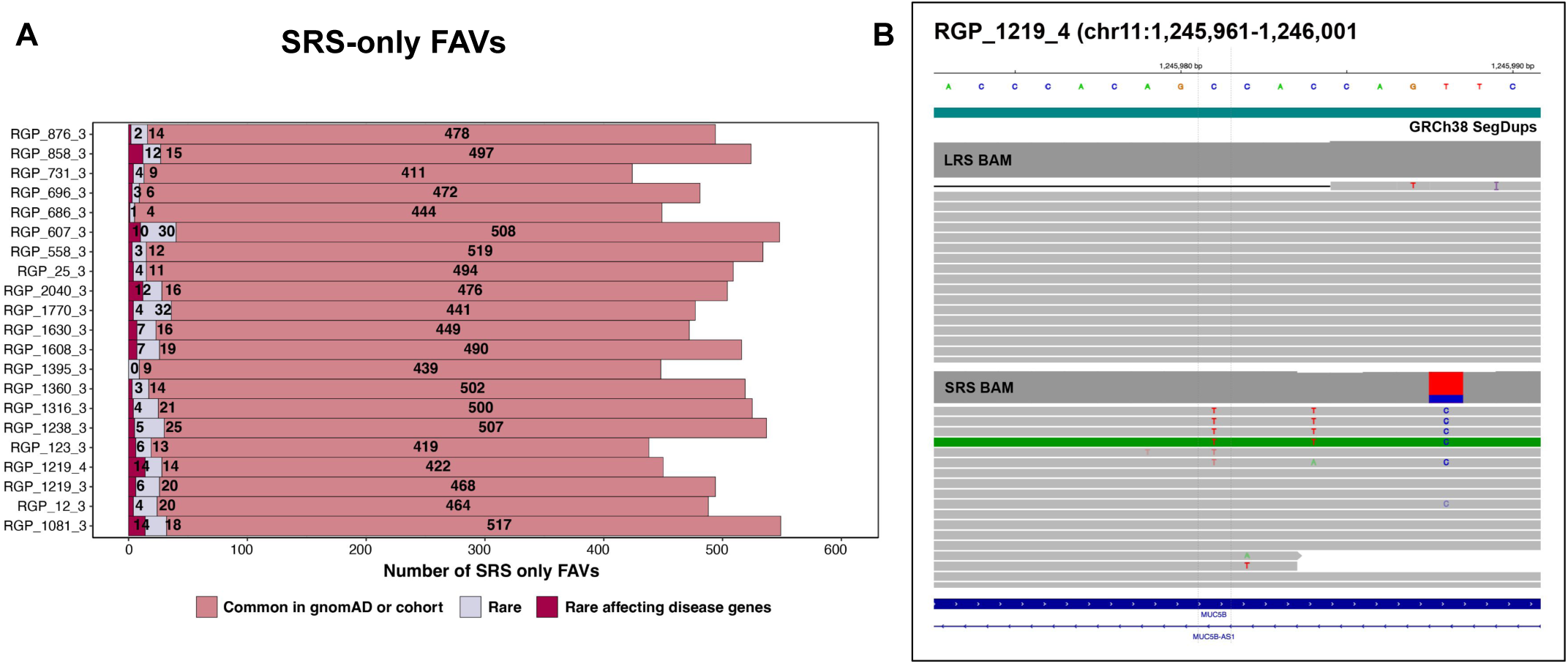
Investigation of SRS-only FAVs on long reads. **A)** A median of 494 SRS-only FAVs per proband. 24 were rare of which 4 were in Mendelian disease genes. **B)** An example of SRS-only FAV (heterozygous C/T) in RGP_1219_4, shows no evidence on LRS, but overlaps annotated segmental duplication in GRCh38. Low quality bases in SRS with other nearby SNVs.

**Supplementary Figure 4:**
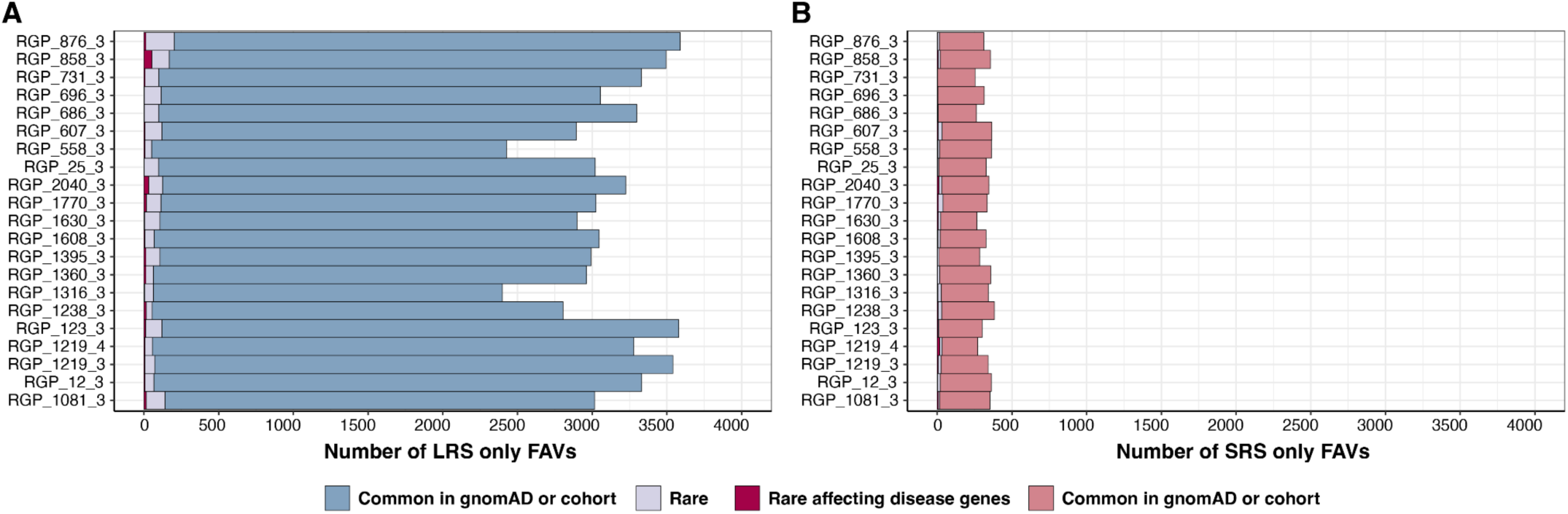
Functionally annotated small variants exclusively called by SRS and LRS (without GQ filtering) **A)** A median of 3046 LRS-only FAVs per proband. 99 were rare, of which 15 were in Mendelian disease genes. **B)** A median of 334 SRS-only FAVs per proband. 22 were rare, of which 5 were in Mendelian disease genes.

**Supplementary Figure 5:**
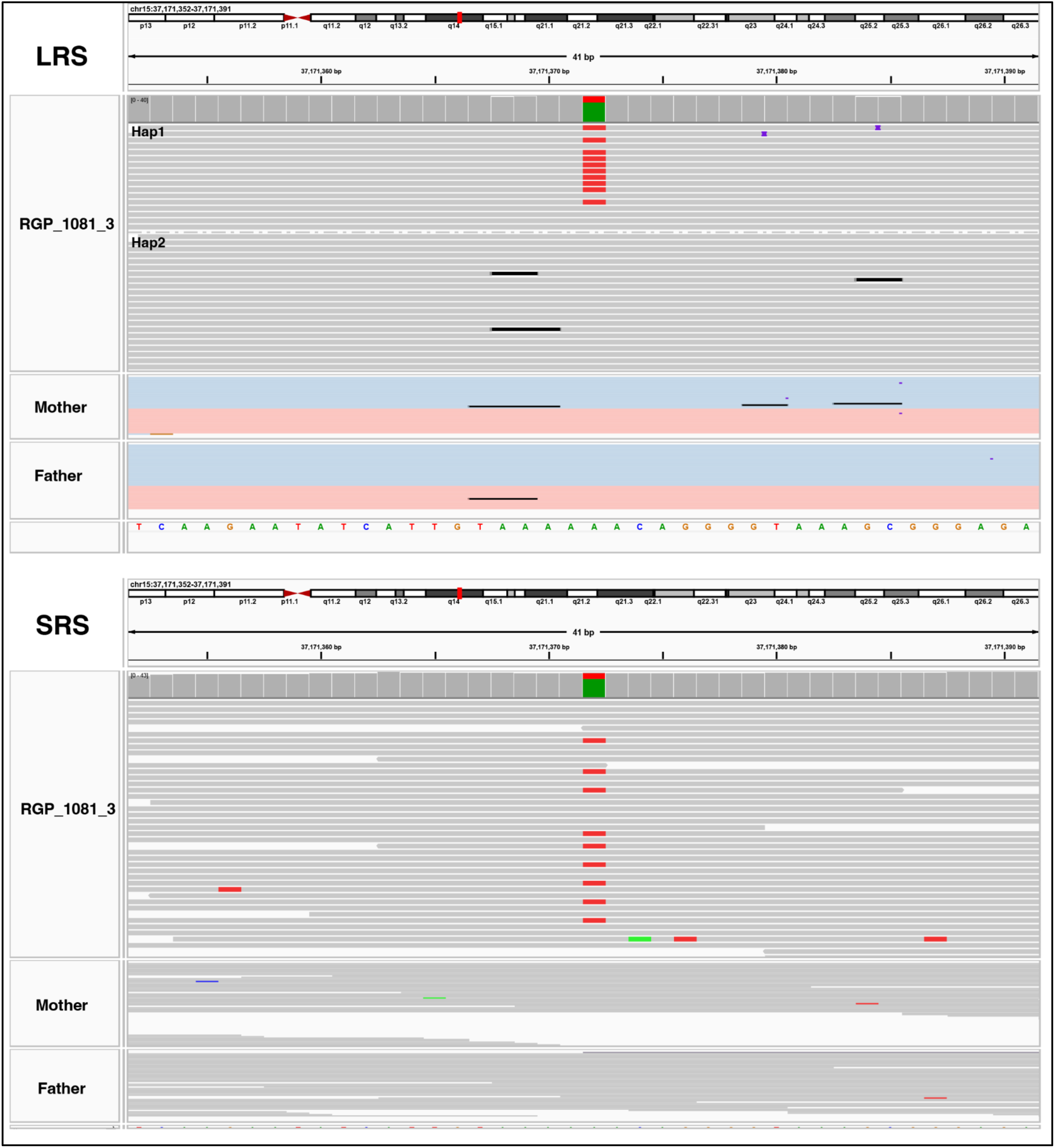
An example of a likely postzygotic mosaic DN-SNV in LRS and SRS reads. In **LRS**, reads are grouped and colored by haplotype. Some reads tagged to haplotype 1 show evidence of the DN-SNV, while others carry the reference allele. All LRS reads have MAPQ60 and high base quality. In **SRS**, the DN-SNV was called but displays a very low AB.

**Supplementary Figure 6:**
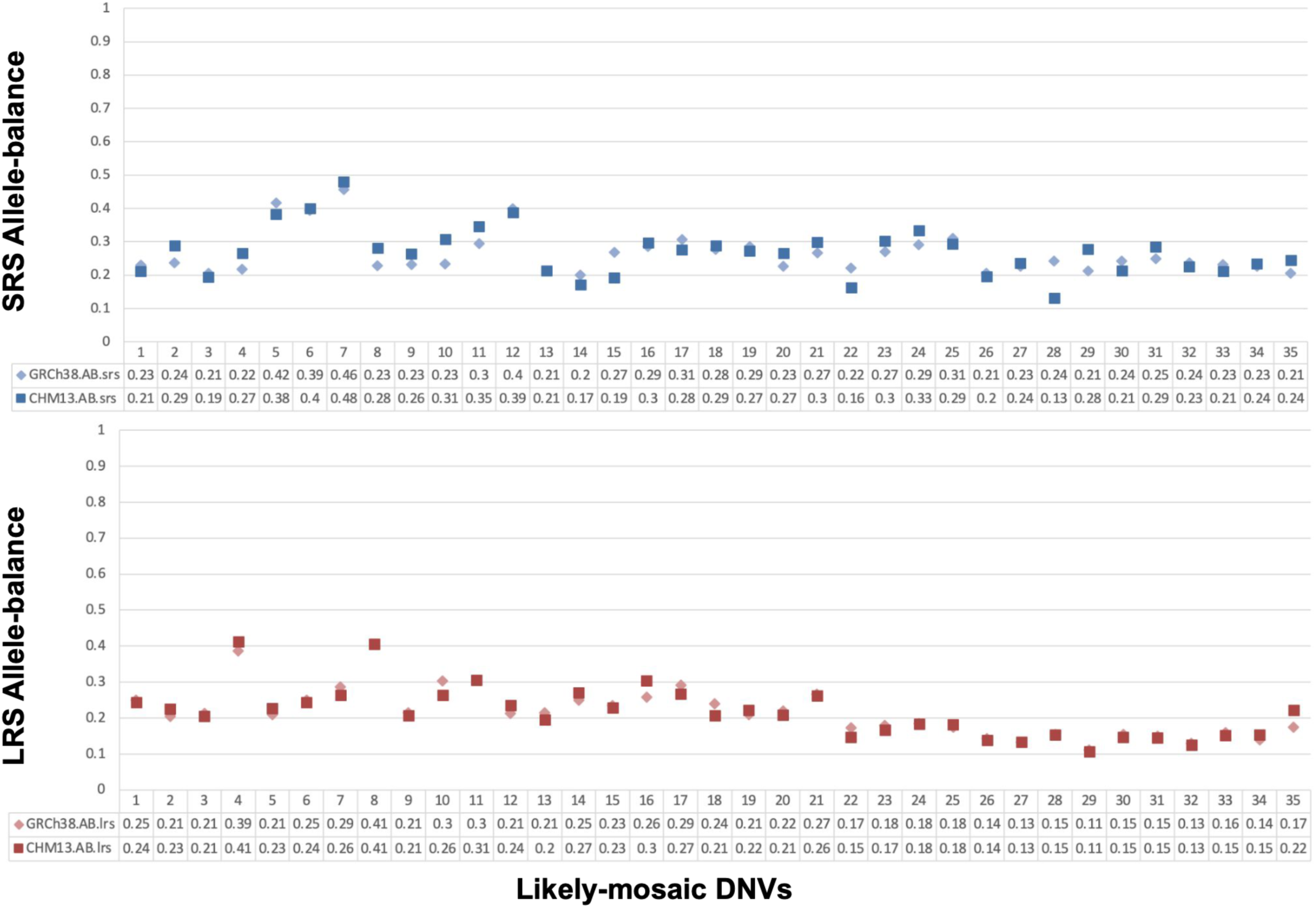
Allele-balance consistent with GRCh38 and T2T-CHM13 mapped short-reads and long-reads. X-axis represents all 35 likely post-zygotic mosaic DN-SNVs. Top panel shows allele-balance of likely post-zygotic mosaic DN-SNVs on SRS, mapped to GRCh38 (light diamond point) and T2T-CHM13 (solid square point). Bottom panel shows allele-balance of likely post-zygotic mosaic DN-SNVs on LRS, mapped to GRCh38 (light diamond point) and T2T-CHM13 (solid square point).

**Supplementary Figure 7:**
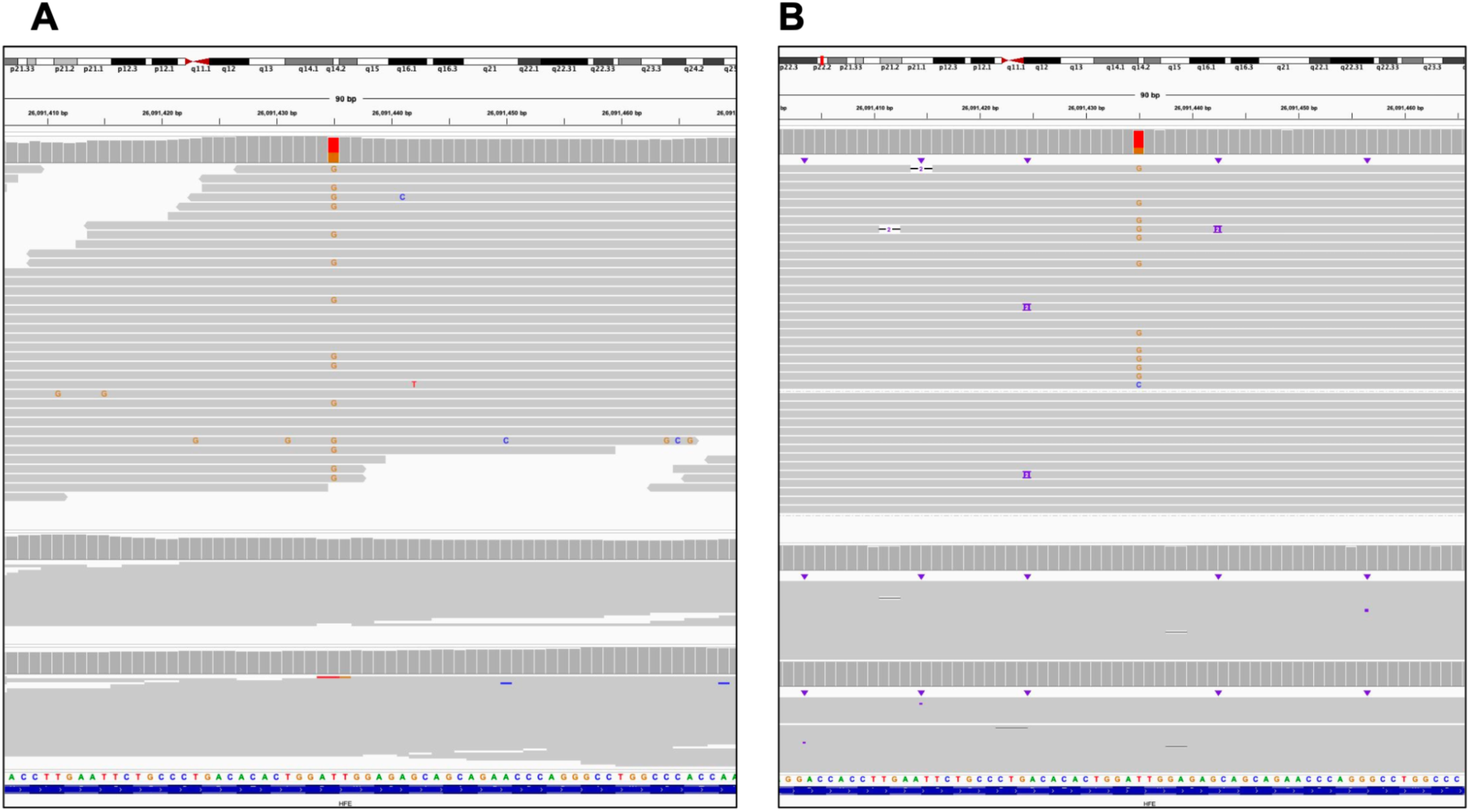
An example of SRS-only likely true-positive DN-SNV. SRS-only likely true-positive DN-SNV in RGP_1770_3 (chr6:26091435, T>G), which was not called by LRS. There is strong evidence of the variant in **A)** short reads (AB=0.41) and **B)** long reads (AB=0.28). However, the presence of an alternate base C and multiple reads with the reference allele may have caused DeepVariant to miss this call. Long reads are grouped into haplotypes.

**Supplementary Figure 8:**
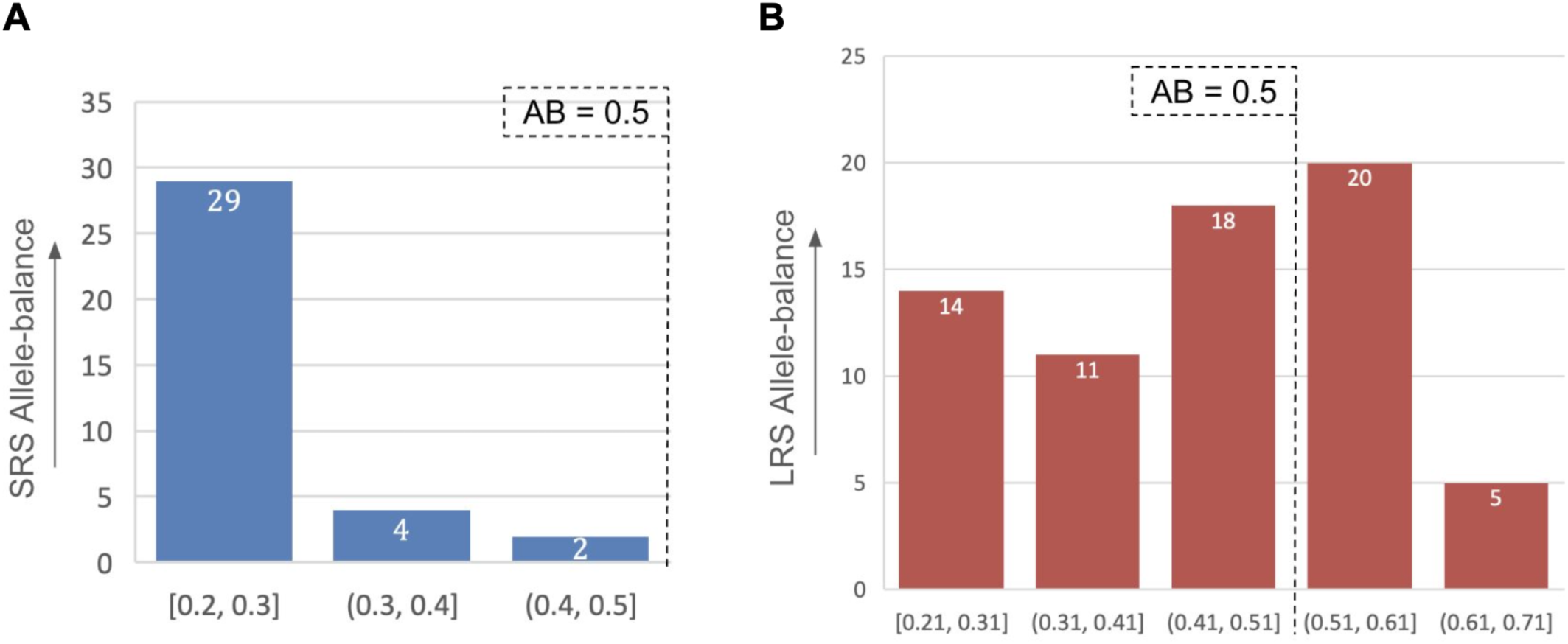
Allele-balance distribution of “likely postzygotic mosaic” DN-SNVs as compared to LRS-only true-positive DN-SNVs. (A) AB distribution of all categorized likely-postzygotic mosaics on SRS is less than 0.5, as compared to **(B)** AB of LRS-only true-positive DN-SNVs, evenly distributed around 0.45.

**Supplementary Figure 9:**
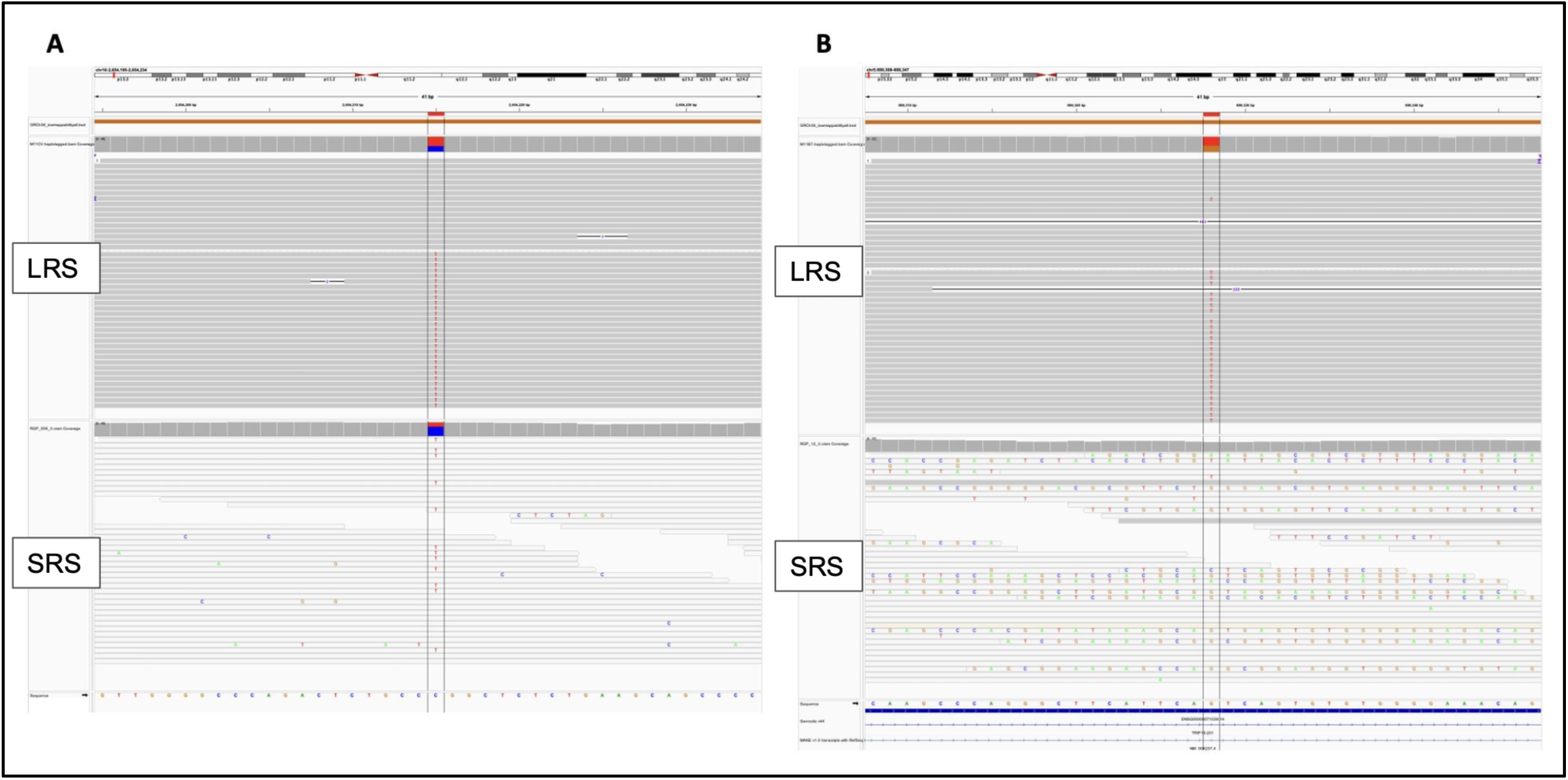
Examples of LRS only likely true-positive DN-SNVs in low short-read mappable regions. The topmost track (Orange) represents GIAB low short-read mappable regions for the GRCh38 reference genome. The second and third tracks represent long read and short read alignments for the proband sample, respectively. **A)** Heterozygous DN-SNV in RGP_558 proband at chr16:2654215 (C > T) was found as a likely true-positive DN-SNV by LRS, but not SRS. All long-reads confidently map with high mapping quality, and the variant is accurately phased. No short read confidently maps to the region (illustrated by transparent mapping quality 0 reads). **B)** Heterozygous DN-SNV in RGP_12 proband at chr16:899328 (G > T) was found as a likely true-positive DN-SNV by LRS but not SRS. It is a downstream variant of gene TRIP13 and affects four transcripts. Long-reads confidently map with high mapping quality, and the variant is accurately phased. Short reads ambiguously map to the region.

**Supplementary Figure 10:**
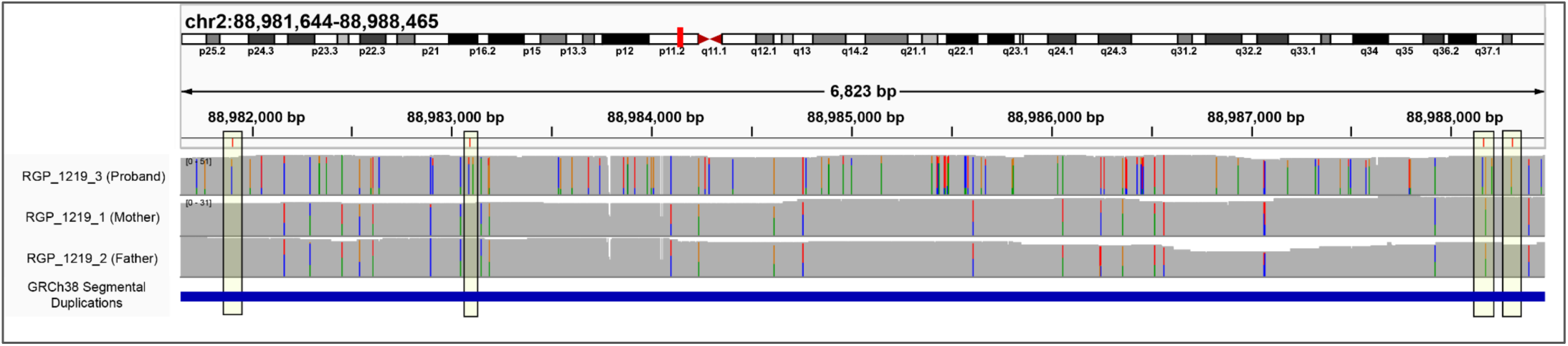
Example of LRS-only likely true-positive DN-SNV clusters on specific chromosomes in the same individual. The first three tracks display the long-read alignment coverage and mismatches for the proband and both parents. Highlighted boxes mark the locations of de novo variants identified in the proband. The blue track at the bottom indicates the GIAB segmental duplication regions for the GRCh38 reference genome.

**Supplementary Figure 11:**
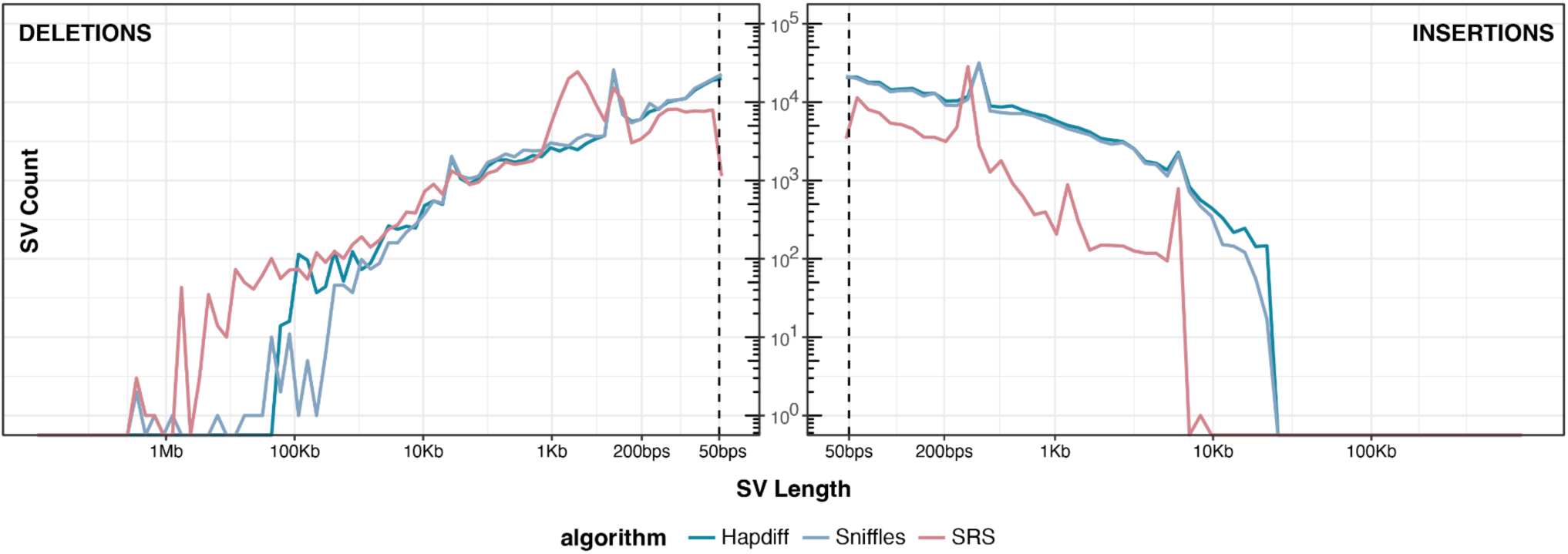
Structural variants length spectrum. Length distribution of deletions and insertions detected by each technology on a log-log axis. Dashed line represents 50bp, the threshold for calling an indel an SV.

**Supplementary Figure 12:**
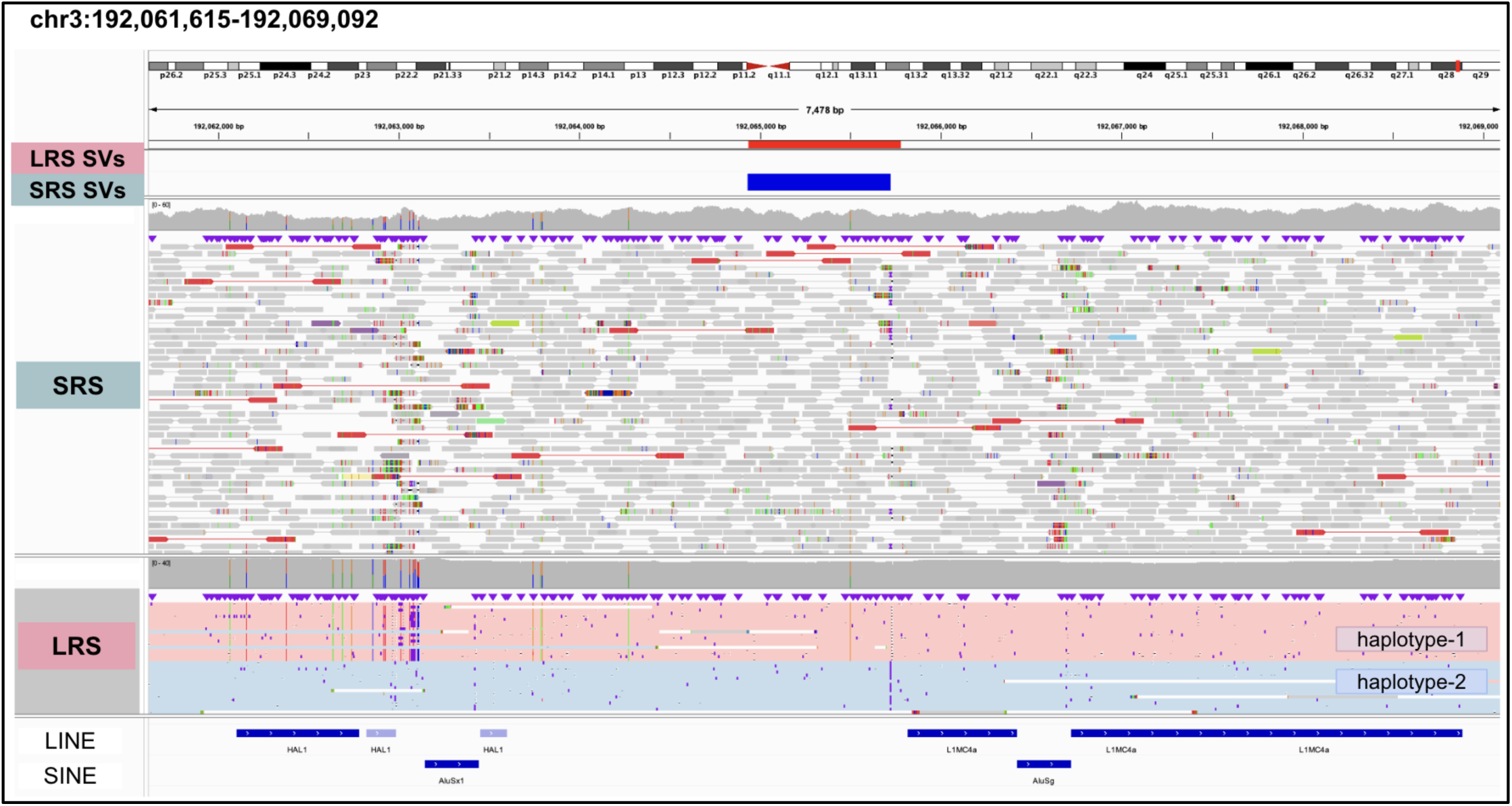
An example of a SRS-only deletion SV that was uniformly covered with LRS. A false-positive 794 bp deletion in RGP_696_3 was called with SRS, unsupported by LRS, showing uniform coverage throughout the region. On the SRS tracks, paired-reads are represented, with red colored pairs meaning longer insert size than expected. The paired-end reads don’t span the region to accurately define deletion boundaries. On the LRS track, reads are colored by haplotype.

**Supplementary Figure 13:**
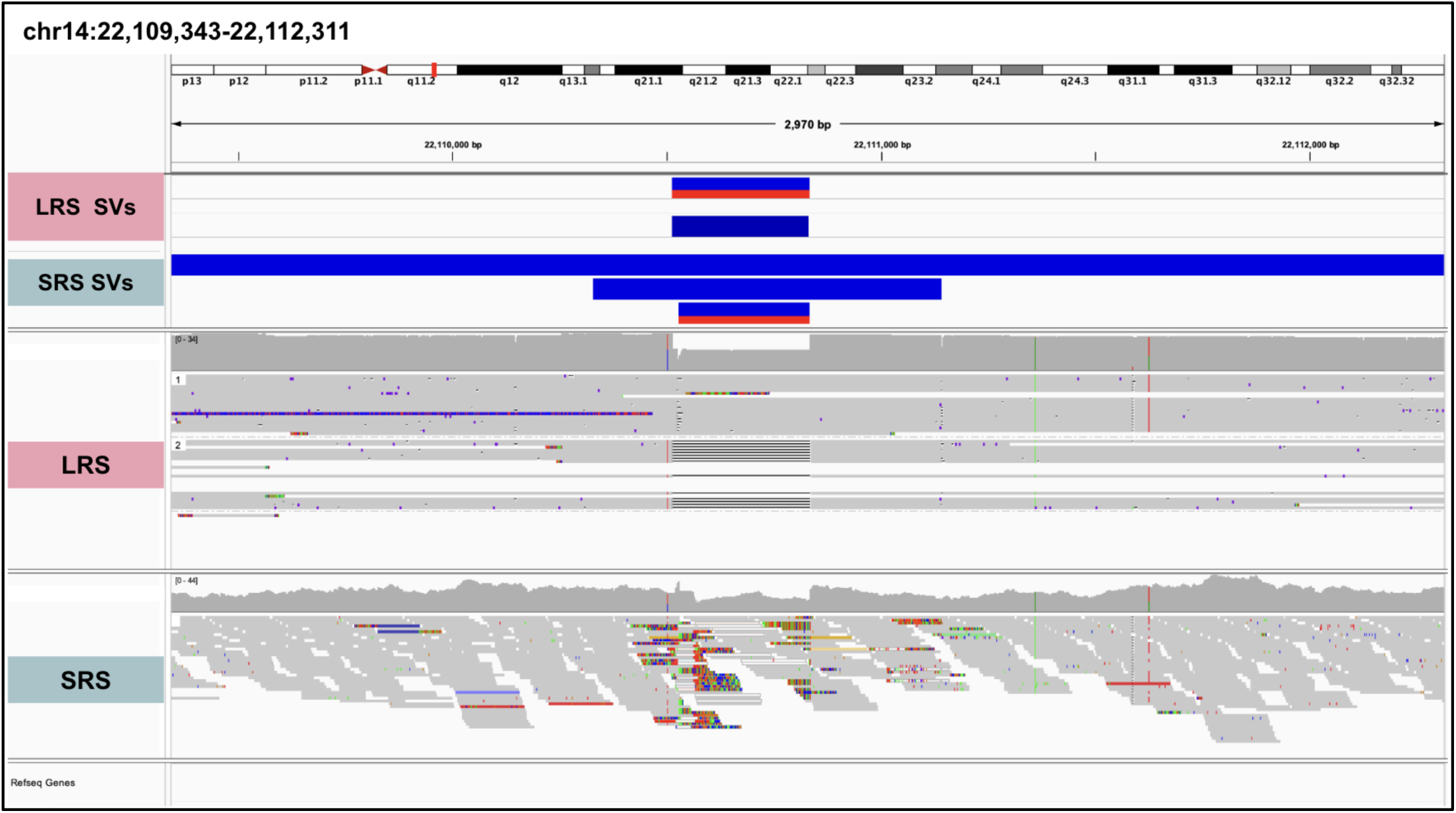
An example of a multiple overlapping SRS-only SVs called versus a single sequence and haplotype-resolved LRS SV. Three overlapping SRS SVs were identified in the region chr14:22,109,343-22,112,311 in RGP_696_3. Two matched the corresponding LRS SV (only after setting a very low overlap fraction of 0.1), while the other (488 Kbp deletion) was categorized as SRS-only. However, this SRS-only SV had very low variant allele balance, and SRS reads did not provide clear evidence for the deletion.

**Supplementary Figure 14:**
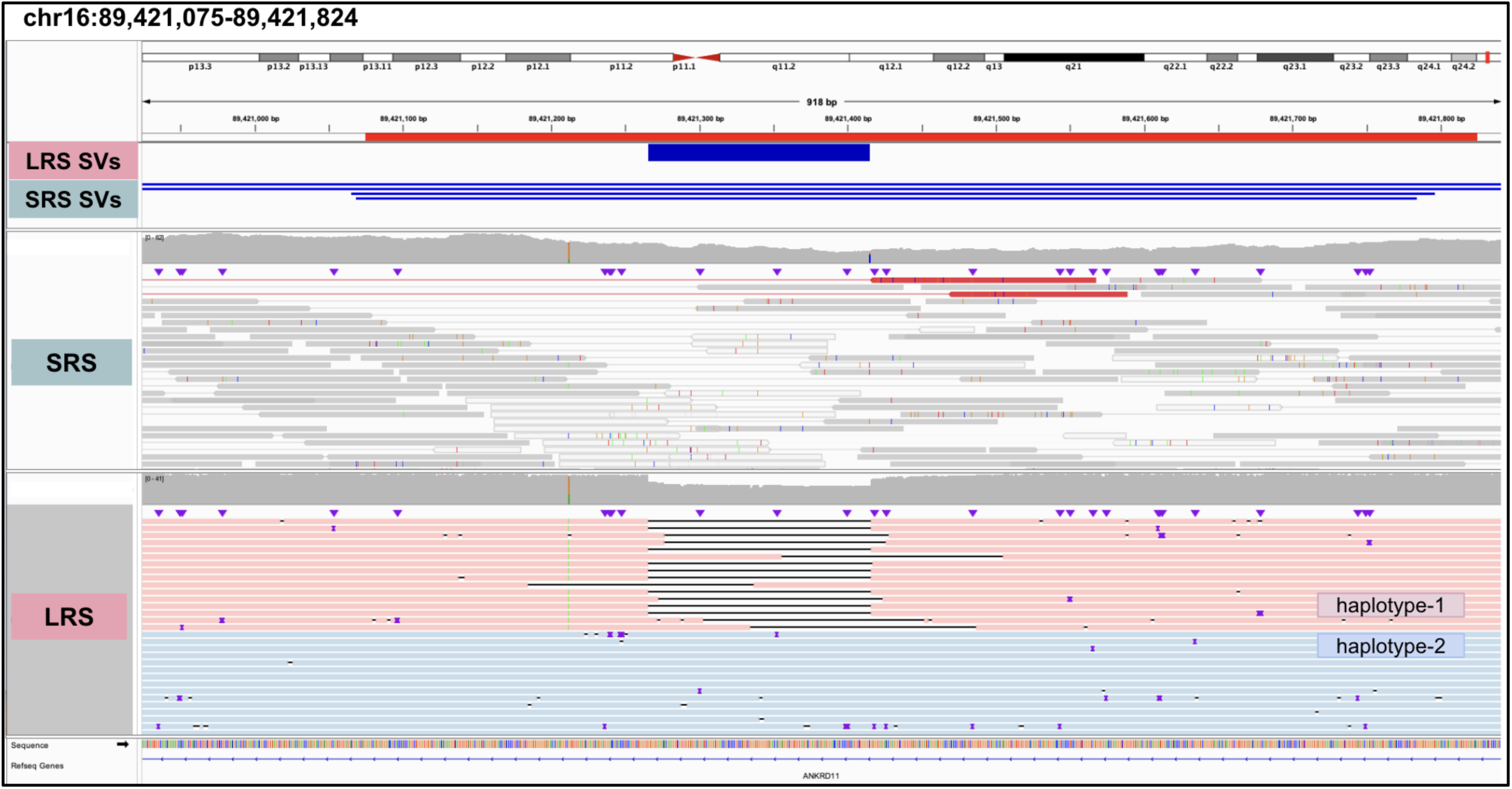
An example of LRS-only deletion SV supported by reads. A 150 bp deletion in RGP_696_3 was called as LRS-only was confirmed by evidence on haplotype-resolved reads. Clearly, in SRS, there is no clear evidence of the deletion breakpoints, and SRS instead calls multiple low-quality SVs in the region.

**Supplementary Figure 15:**
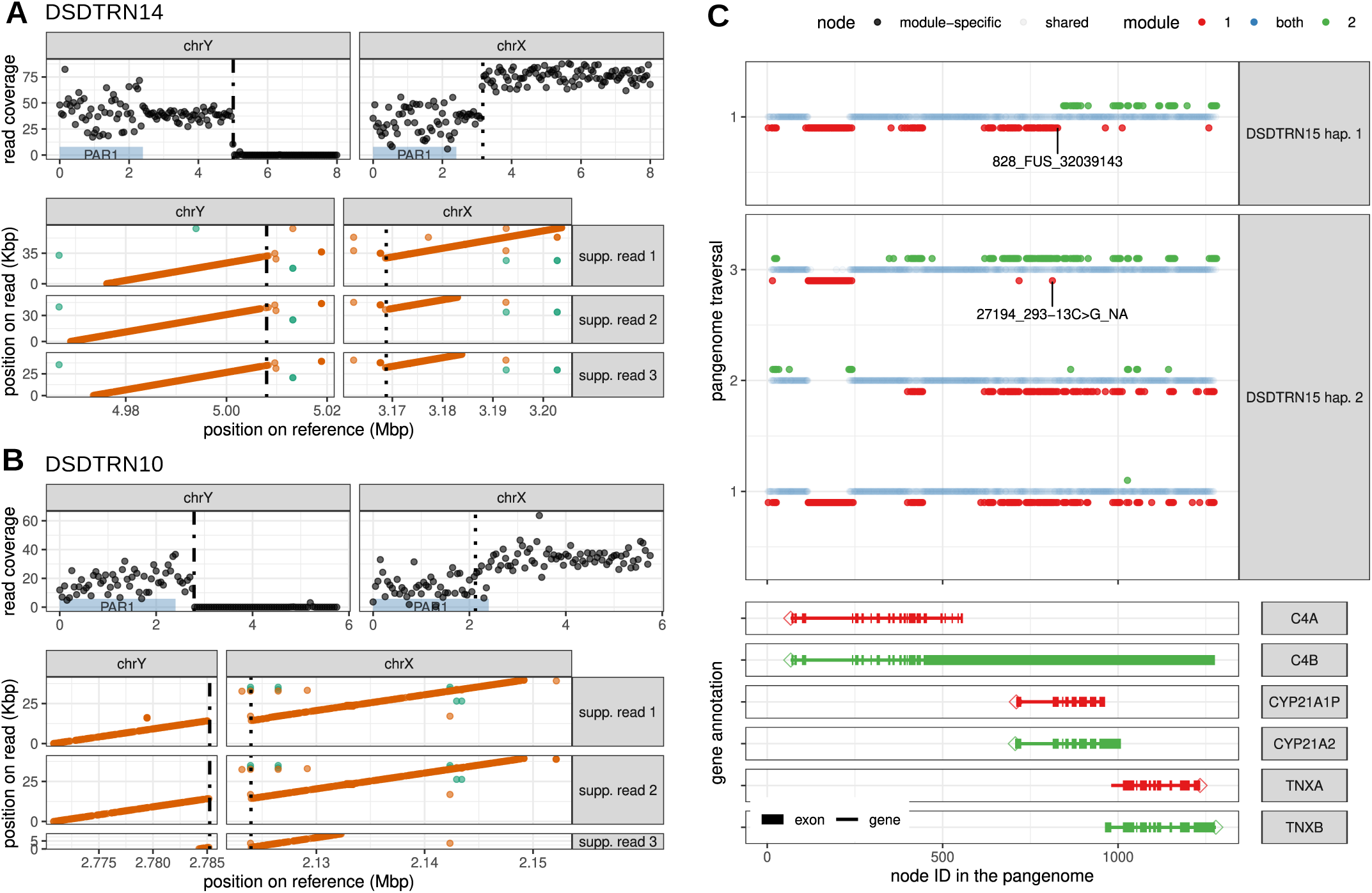
Base-level characterization of complex structural variants. **A-B)** Y-to-X translocations in two probands. The read coverage (top panels) shows one-copy of a small portion of chrY and the loss of one copy of the beginning of chrX. The breakpoints (dotted vertical lines) are consistent with the location where the alignment of the reads breaks, as shown in the dotplots (bottom panels). **C)** *CYP21A2* results for a proband with congenital adrenal hyperplasia. The top two panels represent how each predicted haplotype traverses the pangenome. Each dot represents a node in the collapsed pangenome, colored in red/green if they tend to be specific to module 1/2 in the reference genomes (GRCh38 + HPRC). A fusion between the *CYP21A1P* pseudogene and the CYP21A2 gene is visible as a switch from module 1 (red) to module 2 (green) in the first haplotype. The other pathogenic variant labeled is a “module 1” node isolated within module 2, i.e. a small region of the gene that was likely converted to the pseudogene’s alleles. The bottom panels show the location of the genes in the region.

**Supplementary Figure 16:**
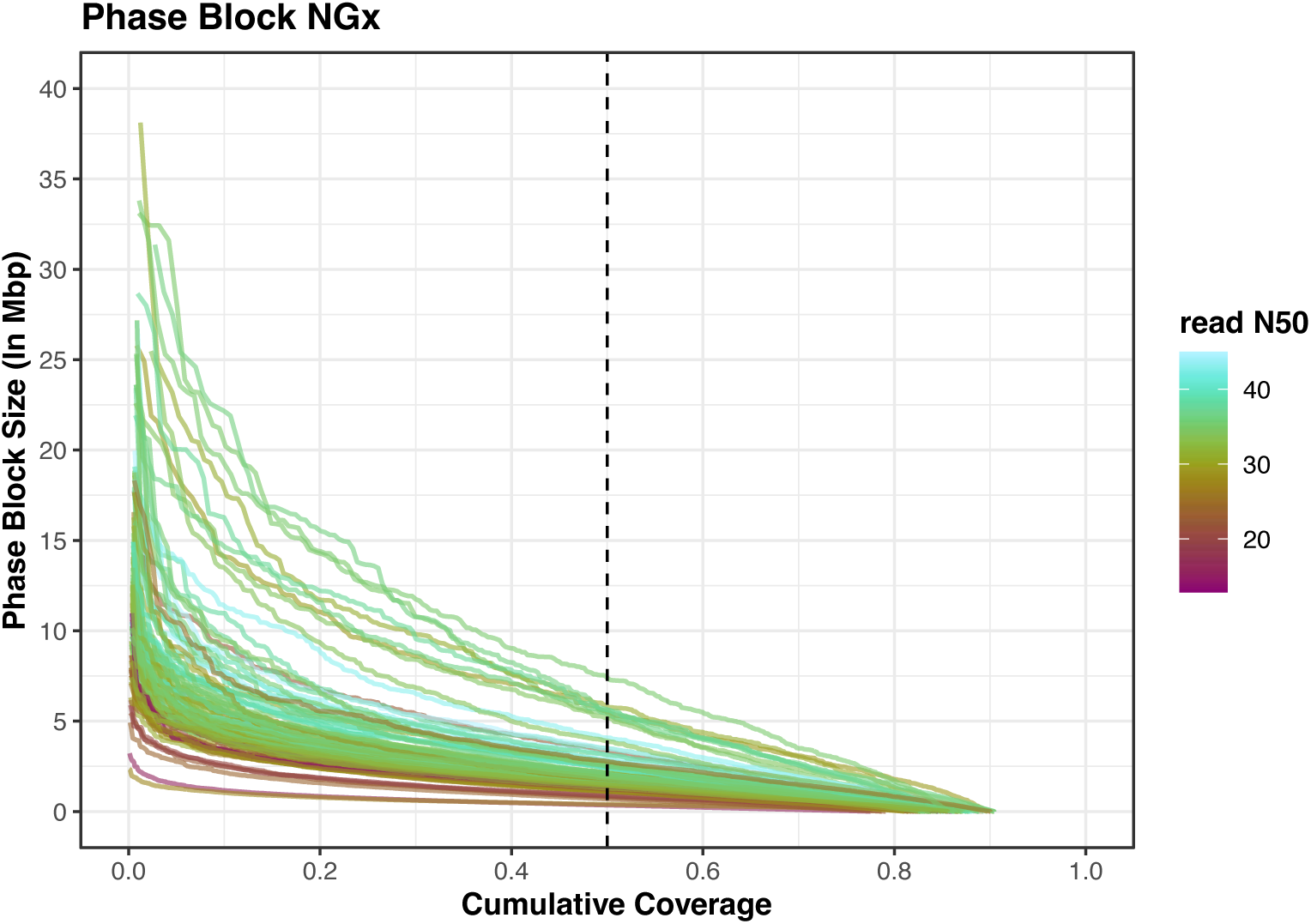
Phase block NG50 statistics. The plot shows the phase block NGx, inferred from harmonized phased VCF. Each line corresponds to a sample, colored by read N50s.

**Supplementary Figure 17:**
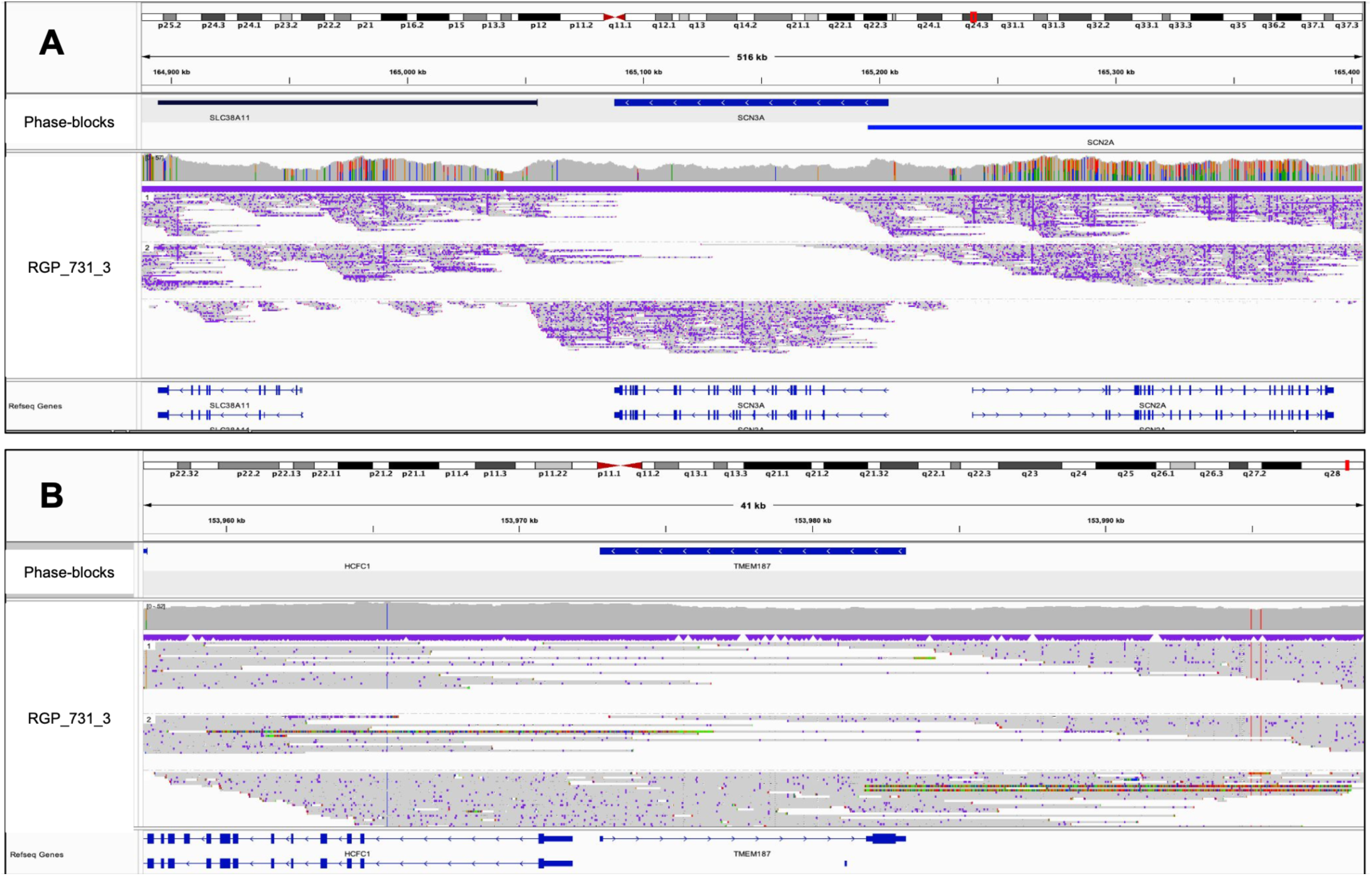
Explanation for some genes overlapped by 0 phase-blocks. **A-B)** Most of the 0 phase-block genes included small or single-exon genes where no heterozygous variants were called, despite sufficient coverage, resulting in overall smaller computed phase-block sizes than possible.

**Supplementary Figure 18:**
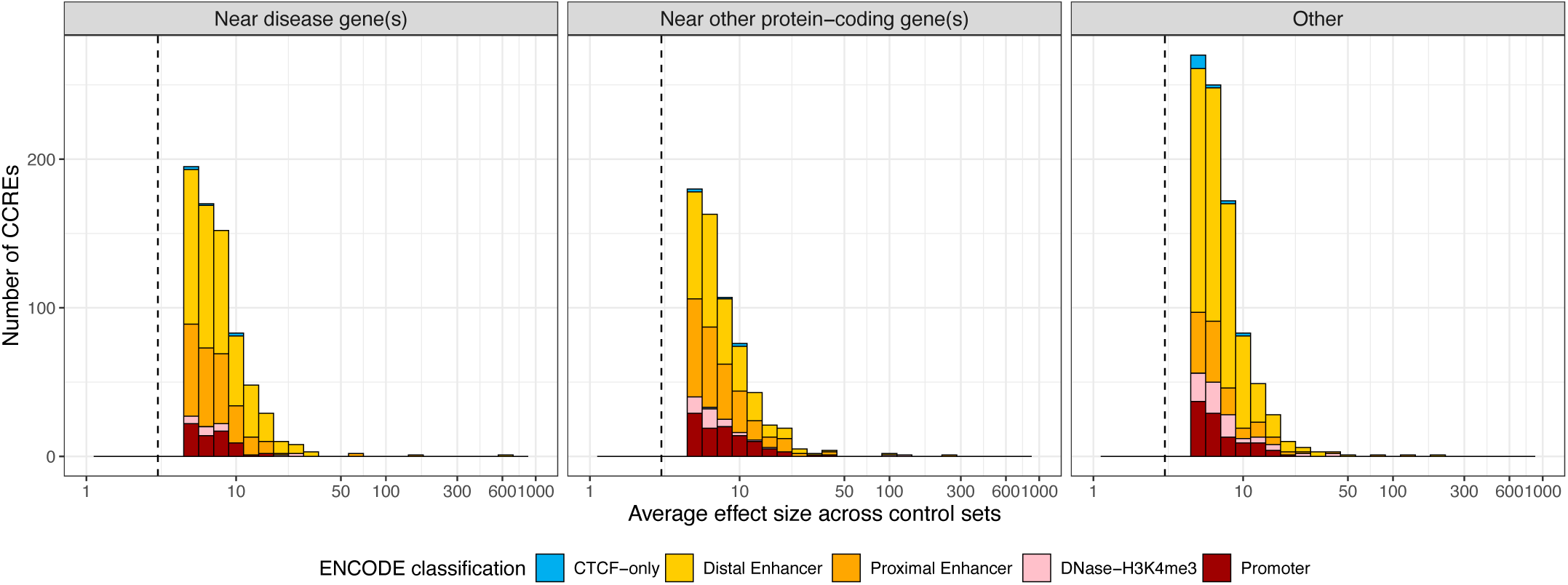
Differentially methylated ENCODE CCREs. Haplotype-specific DM-CCREs distributed by average Cohen’s d across control sets in all probands. The vertical dotted line highlights average Cohen’s d cutoff for significant DM-CCREs per proband haplotype, computed from the methylation profile of controls (control set 1) and remaining probands (control set 2). DM-CCREs are separated into 3 categories (nearness to known disease gene(s) (within ± 1 Mbp), nearness to other protein-coding gene(s) or > 1 Mbps away from any protein-coding gene). DM-CCREs are colored by ENCODE classification.

**Supplementary Figure 19:**
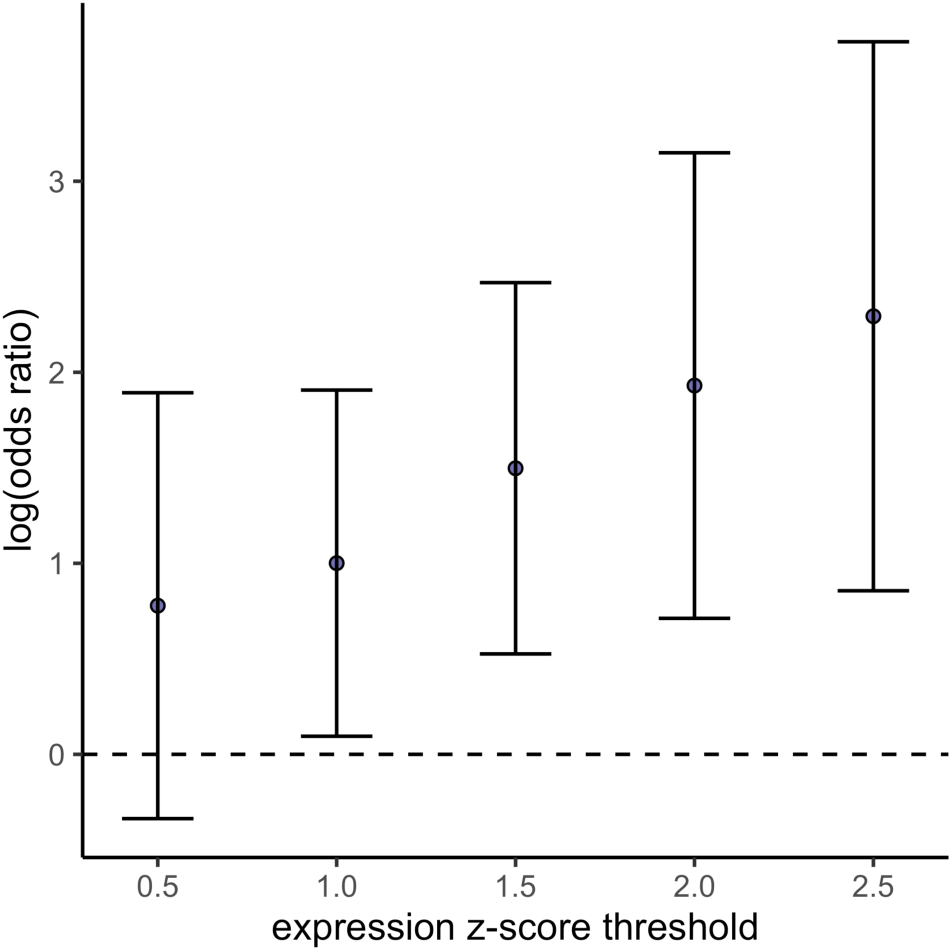
Expression v/s methylation association. Expression v/s methylation association revealed unique DMRs to be associated with nearby changes in gene expression. DMRs were annotated with all genes expressed in blood falling within a surrounding 10 kb window. For these genes, association (y-axis) between DMR status and expression outlier status across a series of absolute z-score thresholds (x-axis) was assessed by logistic regression in 21 affected individuals.

**Supplementary Figure 20:**
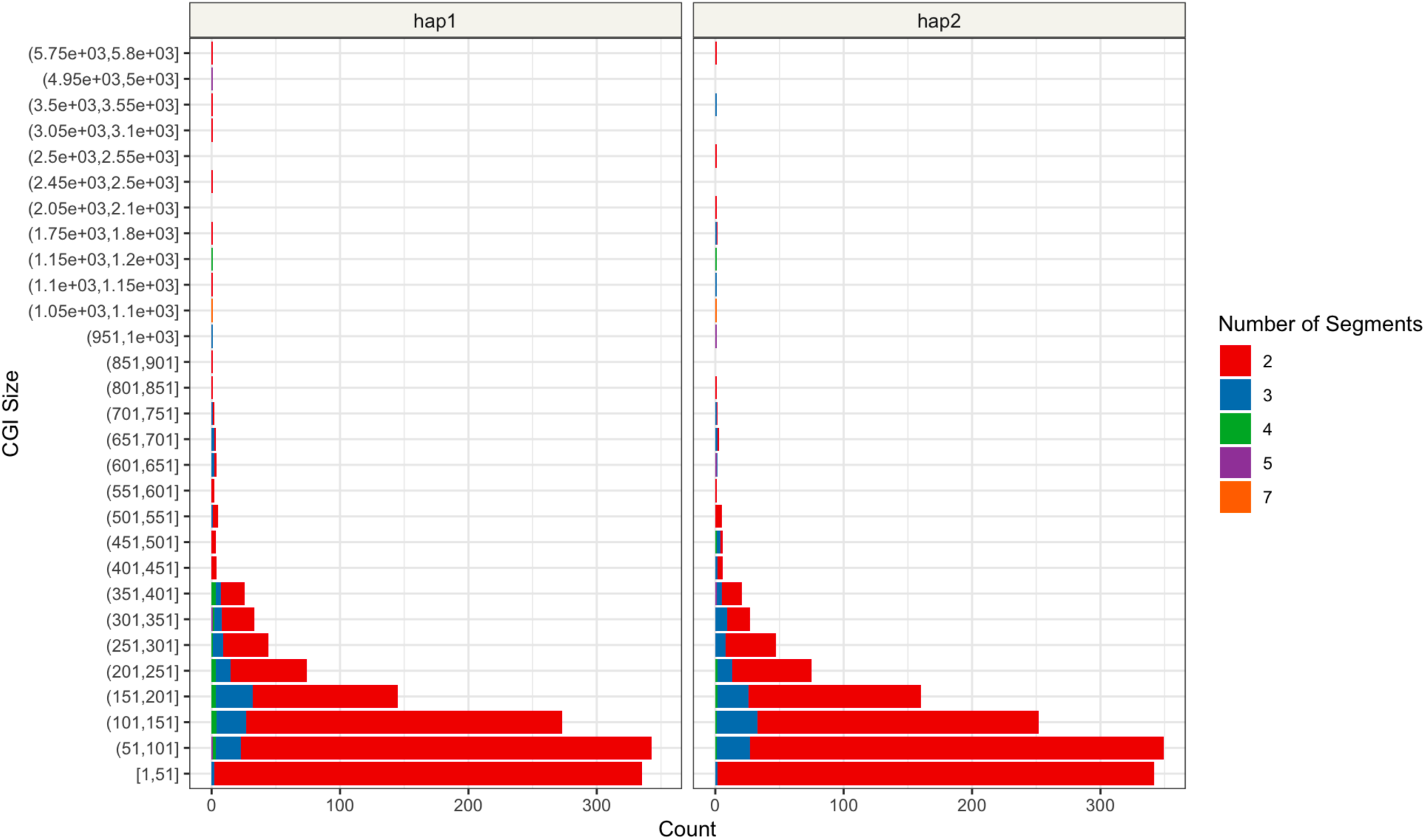
Methylation-based segmentation of CGIs. Using SegMeth on a proband sample, more than 1300 CGIs showed variable methylation patterns, i.e. could be broken into >=2 statistically different segments based on individual CpG methylation.

